# Role of the exercise professional in metabolic and bariatric surgery

**DOI:** 10.1101/2023.04.20.23288698

**Authors:** Matthew A. Stults-Kolehmainen, Dale S. Bond, Laura A. Richardson, Louisa Y. Herring, Bethany Mulone, Carol Ewing Garber, John Morton, Saber Ghiassi, Andrew J. Duffy, Ethan Balk, Charles J. Abolt, Matt C. Howard, Garrett I. Ash, Susannah Williamson, Emilian Rejane Marcon, Melissa De Los Santos, Samantha Bond, Janet Huehls, Osama Alowaish, Nina Brojan Heyman, Bruno Gualano

## Abstract

**Background:** Physical activity (PA) is important for the long-term health and weight management of patients who undergo metabolic and bariatric surgery (MBS). However, the roles of exercise professionals in MBS settings have not been systematically determined.

**Objectives:** To investigate: (1) who are the professionals implementing PA programming in MBS clinical settings; and (2) what patient-centric tasks do they perform?

**Setting:** Clinical and academic exercise settings worldwide.

**Methods:** This multimethod study included a scoping review of PA programs in MBS described in the research literature. Data about job tasks were extracted and provided to 10 experts to sort into categories. Cluster analysis was utilized to find the hierarchical structure of tasks. A Delphi process was used to agree on a final model.

**Results:** The majority of PA professionals were exercise physiologists in the USA and physiotherapists or other types of exercise professionals elsewhere. Forty-three tasks were identified, the most reported being: supervision of exercise, fitness testing, and exercise prescription. Seven higher-order categories were determined: (1) Exercise-related health assessment, (2) Body composition and physical fitness assessment, (3) Lifestyle physical activity and sedentary behavior assessment, (4) Education, instruction, and prescription, (5) Exercise monitoring, (6) Behavioral counseling and psychosocial support, and (7) Dietary support. The following statements were rated an average of 9.0, classifying them as “imperative”: 1) “Pre- and post-operative PA/exercise guidelines for MBS patients are needed”, 2) “MBS programs need to include PA/exercise as part of multidisciplinary care”.

**Conclusions:** The expert group reached a consensus on 7 major classifications of job tasks for the exercise professional. It is important for governing medical associations across the world to formally recognize experienced exercise professionals as playing pivotal roles in continuing, multidisciplinary care for MBS patients. These findings also provide evidence-based information in the effort to solidify these positions within the greater context of healthcare.

**Highlights:**

1. Results identified 43 patient-centered job tasks, which divided into 7 major categories.
2. The most common tasks were exercise prescription, supervision, and fitness testing.
3. Job tasks performed varied marginally by the type of exercise professional.
4. Including exercise in MBS patient care was deemed “imperative” by the Expert Group.

## Introduction

Metabolic and bariatric surgery (MBS) yields profound weight loss and health improvements, although there is substantial variability in the amount and trajectories of these outcomes ^(1)^. Some of this variability may be attributed to the physical activity behaviors of patients who had MBS. In this population, more moderate-to-vigorous intensity PA, less sedentary behavior, and more steps are associated with greater weight loss, better weight maintenance, better bone health, and improvements in cardiometabolic health parameters ^(2–6)^. However, patients overall do not become spontaneously active after surgery. Indeed, an average patient is insufficiently active before surgery and makes only small changes in activity behaviors after surgery, thus remaining insufficiently active ^(7)^. In the absence of exercise, several important benefits of MBS (e.g., improvements in insulin resistance, inflammatory markers, endothelial function) are reversed, even in a short period of time, as has been demonstrated in a recent trial ^(8)^.

The difficulties that patients experience in making substantial changes in their activity behaviors suggest that they need additional information, intervention, and support to make such changes ^(9)^. Indeed, structured exercise and physical activity programming in combination with dietary counseling may help to optimize and sustain MBS outcomes ^(4–6, 10)^. Unfortunately, it is likely that such programs are infrequently provided in real-world, clinical settings ^(9, 11)^. Likewise, there is some data suggesting that even simple exercise guidance is not routinely provided in MBS health services worldwide ^(9, 12)^. This may partly be because while established exercise guidelines currently exist for hypertension, diabetes, and other chronic conditions ^(13)^, there are no specific guidelines for exercise with MBS. Furthermore, effectively translating exercise programs from clinical trials into clinical practice is difficult to achieve ^(3, 14–16)^. Despite these shortcomings, insurance companies often require the provision of exercise counselling as a condition for surgical approval ^(17)^. For instance, the current Aetna policy refers to the exercise physiologist as one professional qualified to supervise a “multicomponent behavioral intervention program”^(18)^.

In the effort to translate physical activity research into clinical practice, Koorts and colleagues ^(19)^ developed the PRACTIS Model. According to this framework, several major considerations must be initially targeted, including the clinical population, context/place, intervention characteristics and the people conducting the implementation – the interventionist. While the first parts have been widely discussed in the literature ^(20)^, the last component has been infrequently addressed across the medical literature ^(16, 21, 22)^. Key questions surrounding exercise personnel include: a) who delivers the intervention, b) what skills, education, and qualifications should they hold, and c) how are they trained and supported? While this critical component of the PRACTIS Model has been addressed in primary care ^(22, 23)^, it has not received adequate attention in the context of MBS. Typically, this is not the physician or surgeon, who usually lacks the time and training ^(24)^, but it may include an exercise professional (EP) working as part of a multidisciplinary team ^(25)^.

Exercise professionals comprise a large and diverse group of individuals all aiming to improve health of patients through physical activity ^(26)^. Unfortunately, only recently have guidelines been established to report this information in the scholarly literature ^(27)^, and researchers are just beginning to conform to this initiative ^(28)^. To fill these important knowledge gaps, the major objective of the current study was to utilize a rigorous multimethod approach ^(29)^, including a scoping review and thematic analysis, to answer two main questions: 1) Who are the exercise professionals conducting PA programming in MBS? (e.g., exercise physiologists vs physiotherapists/physical therapists), and 2) What are the prominent clinical exercise job tasks conducted by EPs in an MBS setting? In this effort, we secondarily aimed to describe: a) the qualifications, training, and/or background of the professionals who are delivering the interventions and b) if skills of the exercise professional varied by field. Lastly, we aimed to address issues pertinent to the implementation of PA programming in MBS (e.g., guidelines, reimbursement).

## Methods

To appropriately address the research questions, we utilized a blend of qualitative and quantitative methods – The Systematic Clinical/Organizational Role Evaluation (SCORE) technique, which integrates the: AGREE model ^(30)^, inductive thematic analysis ^(31)^, Card sort (Q) analysis ^(32)^, Hierarchical cluster analysis ^(33)^, and the Delphi process ^(34)^. The AGREE Model for clinical decision making, which has been used for other exercise clinical decisions, like the Physical Activity Readiness-Questionnaire (PAR-Q) ^(35)^, was modified for the current task. See Figure 1.

**Figure 1.**
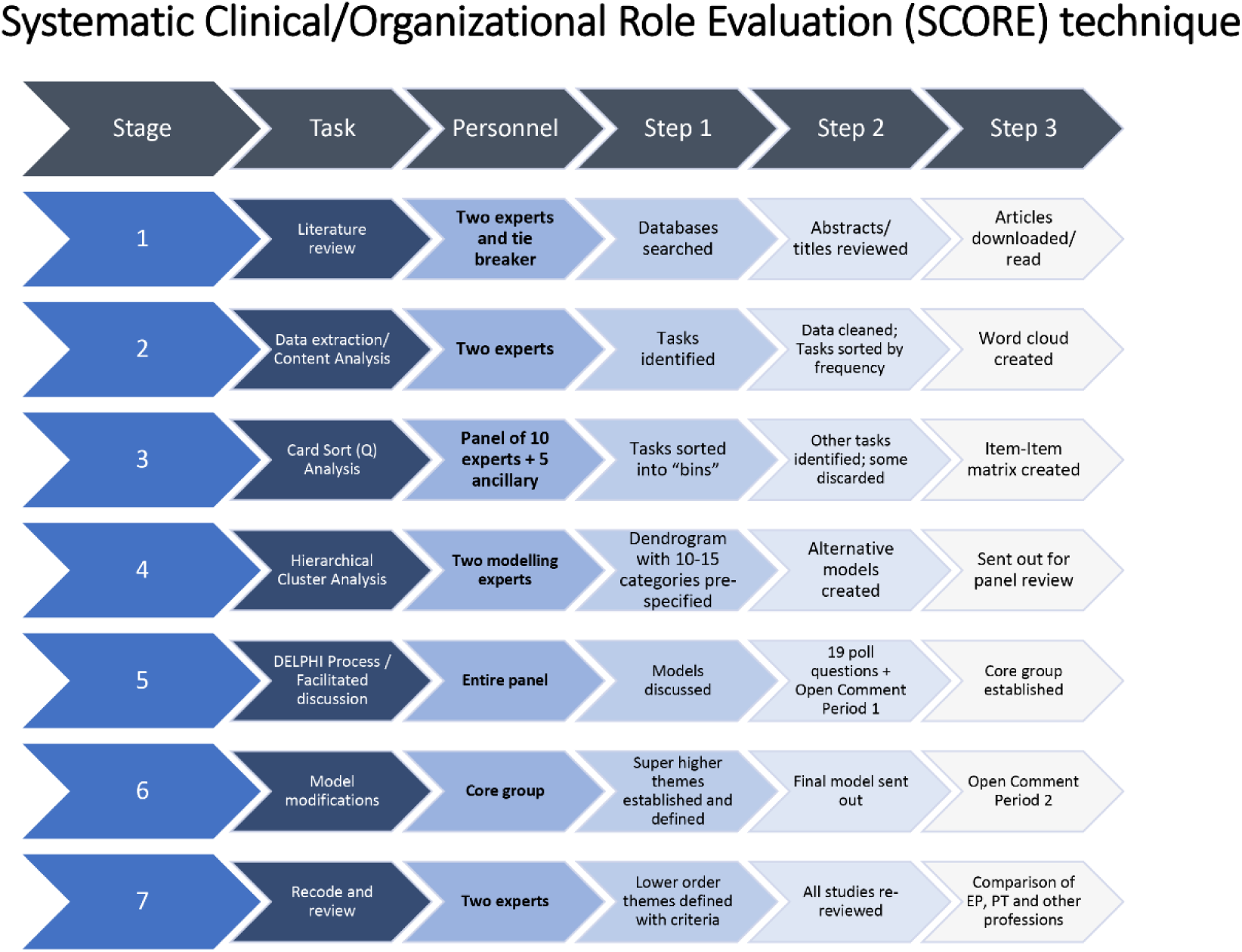
Systematic Clinical/Organizational Role Evaluation (SCORE) technique used in the current study

### Literature review

Per the AGREE model ^(30)^, we started with a scoping review of the literature (Stage 1) that took place between June, 2021 and January, 2022 in the Web of Science (Core Collection) and PubMed databases. PRISMA guidelines were followed ^(36)^. The review included peri-operative MBS literature from the United States and globally to capture a wider lens of therapeutic exercise interventions. Titles and abstracts were searched, with the first term being “bariatric” and the second term being “exercise” or “physical activity”. Database results were filtered by language (English and Portuguese only), use of human subjects and studies publishing original data (e.g., eliminating comments, reviews, guidelines, editorials). Duplicate studies were then eliminated. During screening of titles and abstracts, studies were required to describe exercise and physical activity training and/or education programs (e.g., not just observe participants’ PA over time). Refer to Supplement 1. Some articles were obtained by scanning: a) review papers, b) personal archives (i.e., relevant articles already in authors’ possession), and c) reference lists from all studies obtained, and d) Google Scholar alerts for “Bariatric” and “exercise”. Full text articles were obtained and read for eligibility. See the PRISMA checklist in Supplement 2.

**Supplement 1.**
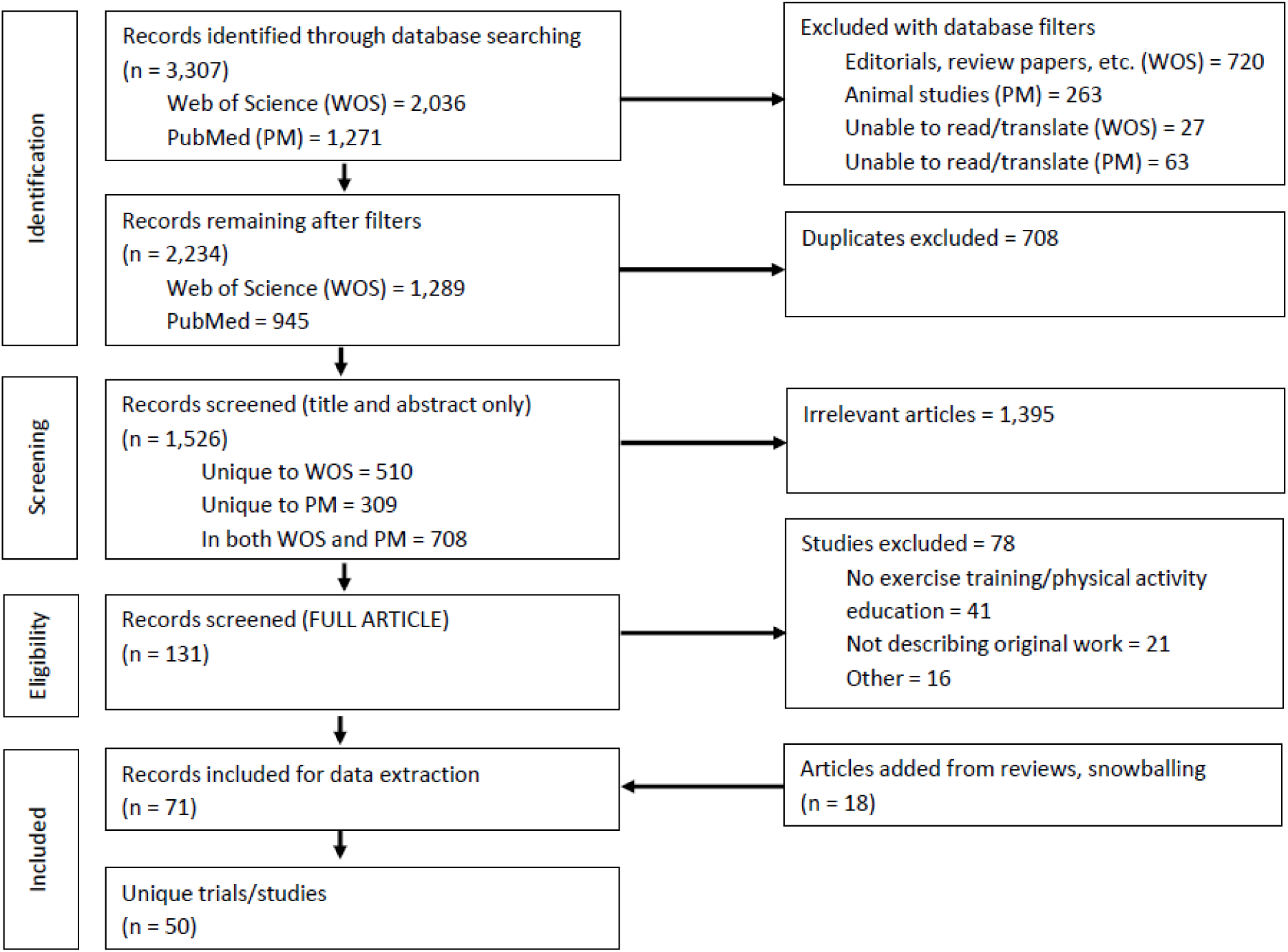
Prisma CONSORT diagram demonstrating the literating the literature search.

### Data extraction

Two investigators (MSK and MD) extracted data related to patient-centered (clinical) exercise and physical activity program tasks completed by the exercise professional (Stage 2). An initial list of job tasks was identified through methods described in Supplement 3.

##### Supplement 3. Data extraction protocol

Two investigators (MSK and MD) performed the data extraction (Stage 2), which first proceeded for information related to the “who” of the exercise program (i.e., type of exercise professional who delivered physical activity programming). Any information regarding education, clinical training, credentialing, and affiliation of the exercise professional was also noted. Gathering this type of data presents some difficulty, because: a. most research articles do not directly state what provider/person completed each aspect of the study, b. authorship lists on most studies represent individuals from diverse academic backgrounds, c. not all study personnel are acknowledged, d. the term used to describe exercise professionals varies across geographical regions, such as: exercise physiologist, physical therapist, exercise specialist, trainer, health educator, etc., and e. missing information due to journal space constraints. Consequently, for studies going back to 2010, the corresponding author in each study’s byline was emailed by the first author with follow up questions.

These same investigators then extracted data related to patient-centered (clinical) exercise and physical activity program tasks completed by the exercise professional. To accomplish this, we decided to use a more inductive approach – relying on the bariatric exercise literature itself and not depending, a priori, on definitions of job tasks as stated by documents from various professional organizations about the role of exercise practitioners (1-4), which may or may not be applicable, or may be inaccurate or insufficient in this setting. By “clinical” or “patient-centered”, it is meant that we did not attempt to focus on tasks related to basic science questions (e.g., laboratory assays), administration, or other activities not directly related to patient contact.

An initial list of job tasks was identified through three means. First, by identifying instances where the role of the exercise professional were directly stated (usually as preceded by strong action verbs) (i.e., created the exercise program, screened patients, completed assessments, supervised/monitored training, provided counseling). Second, in studies conducted entirely by exercise physiologists, one would presume that all activities were conducted by these exercise professionals. Third, a corresponding author declared such actions in a personal communication. Two investigators systematically evaluated and interpreted publications independently to minimize bias (MSK and MD).

##### References

1. American Physical Therapy Association. About Physical Therapists and Physical Therapist Assistants 2023 [Available from: https://www.choosept.com/why-physical-therapy/about-physical-therapists-and-physical-therapist-assistants.
2. American College of Sports Medicine. ACSM Certified Exercise Physiologist: Exam Content Outline 2023 [Available from: https://www.acsm.org/docs/default-source/certification-documents/acsmep_examcontentoutline_2020.pdf?sfvrsn=d413f75_16.
3. Froment FP, Olson KA, Hooper TL, Shaffer SM, Sizer PS, Woodhouse LJ, et al. Large variability found in musculoskeletal physiotherapy scope of practice throughout WCPT and IFOMPT affiliated countries: An international survey. Musculoskeletal Science and Practice. 2019;42:104-19. doi: https://doi.org/10.1016/j.msksp.2019.04.012
4. World Confederation for Physical Therapy. Description of physical therapy: Policy statement. 2019.

## Thematic and Card Sort (Q) Analysis

In the third stage, these data were categorized with a mixed inductive thematic analysis ^(29)^ and card sort (Q) analysis ^(32)^. Data were systematically reduced by MSK and MD, who conducted cleaning and recoding. This included nominalization of verbs. Data were examined: a) first as multiple words (e.g., “heart rate monitoring”) and then, b) connected/adjoined with a hyphen (e.g., “HR-monitoring”) to facilitate visualization. Tasks were then compared for overlap and agreement between investigators. Unique job tasks were entered into a free word cloud program (freewordcloudgenerator.com, Salt Lake City, UT) to visually inspect (black background, rainbow color pattern font), categorize by word frequency, and provide labels ^(37)^. See Supplement 4.

**Supplement 4.**
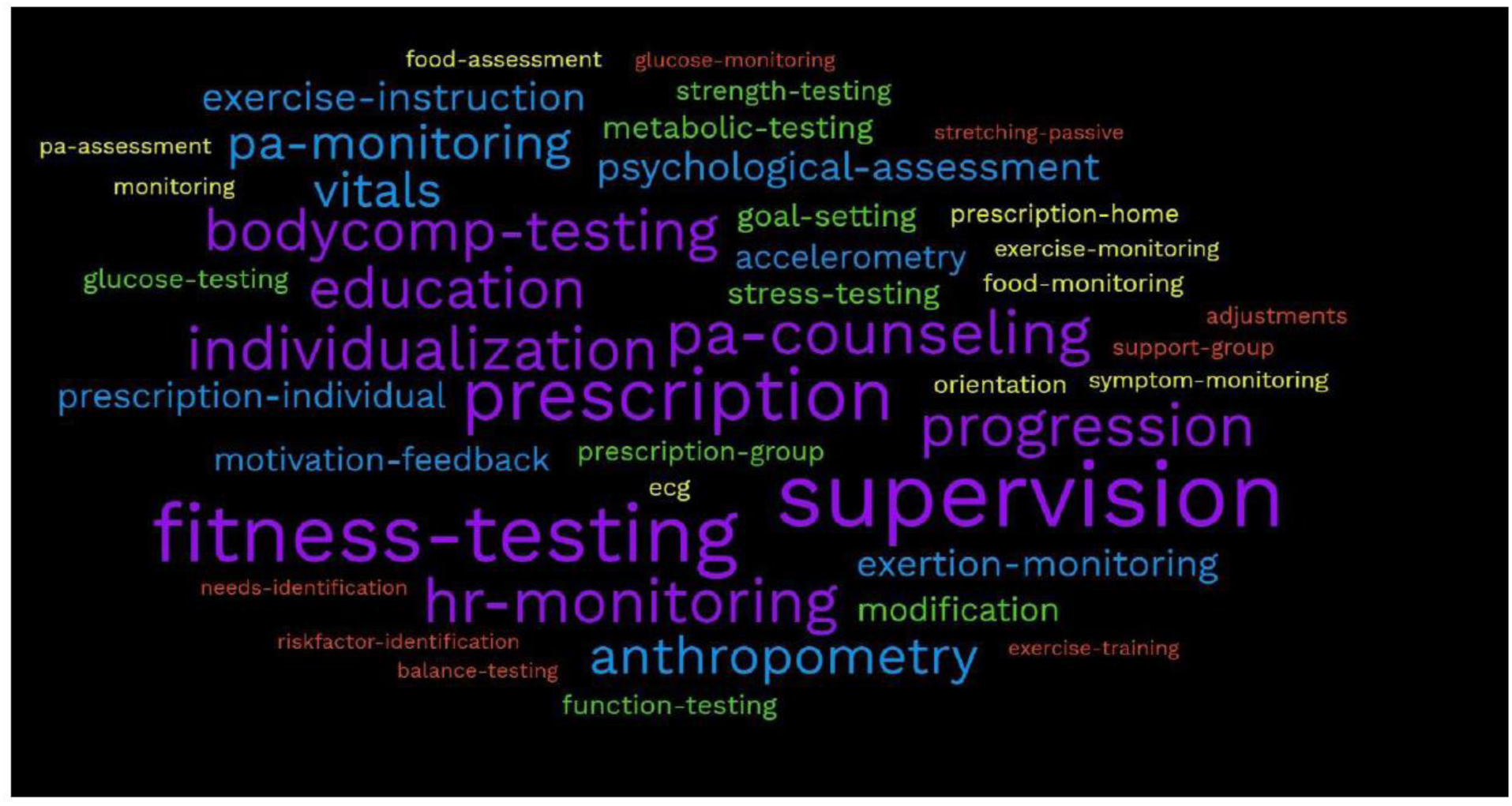
Word clould illustrating common exercise job tasks presented to the research team for the bin sort assignment.

An expert group of bariatric clinicians, exercise practitioners, and scientists were invited to complete the sort task. The Delphi process requires 8-10 people, but due to final exams, summer holidays, we did not expect that all people would respond. Therefore, we over-sampled (15) so at least 8 would be involved at each stage. Ten had complete and valid responses submitted on time. The remainder did not respond in required timeframe (3), had incomplete responses (1) or completed the form erroneously (1). Those analyzed included four exercise physiologists, two registered dietitians (RDs), one advanced practice registered nurse, one surgeon, and two exercise scientists). Four were exercise practitioners in MBS clinics, each with a minimum of 9 years of practice in the area, for a total of 66 years (MSK, LR, JH, RM). Those not responding at this stage continued to follow the process, evaluate data, and more actively participate in later stages. These individuals independently inspected the data, identified related tasks and sorted/categorized these into “bins”/clusters starting with the most frequent job tasks observed. To prevent author word choice from hiding findings (e.g., “body composition testing” vs. “analysis of body composition”), high frequency words were collapsed with similar words and then re-analyzed. The initial goal was 10-15 bins or clusters, based on Hsieh & Shannon ^(31)^. Respondents could elect to discard items or suggest new ones. Responses from these ten experts were included into an Item-by-Item matrix, which was used to determine *distance* of each item from the others ^(32)^.

## Hierarchical Cluster Analysis (HCA)

In Stage 4, a hierarchical cluster analysis was then utilized (Matlab v R2022a) ^(33)^ to create Dendrogram models based on distance between word items. Two alternative Dendrogram models were generated for group consideration. Both lower order and higher order themes were detected from the dendrograms. To identify higher-order themes (HOTs) (to generate the a priori, 10-15 themes) a *distance threshold* was set to 7. To identify 7 super-higher-order themes (SHOTs), a distance threshold was set to 9.

## DELPHI Process/ Issues in bariatric exercise survey

In Stage 5, a process of reaching consensus among the expert group was initiated to determine a final list of job task categories and to address related issues, based on the Delphi Technique ^(34)^. This included 2 group discussion sessions (both in June, 2022) to: a) review the HCA, b) consider adding, deleting and/or merging tasks, c) consider ranking and ordering items, etc. Then a survey developed by a Core Group (MSK, DB, LR and LH) with 19 open and closed questions (in 4 sections) was sent to all respondents. Section 4 included 4 statements about issues in bariatric exercise - rated on a 0 (no importance) to 9 (imperative) scale. An example (the first statement) is, “Pre- and postoperative PA/exercise guidelines for bariatric surgery patients are needed”. Another two weeks were permitted to respond. “Consensus” was defined by > 80% agreement for the first 3 sections of questions and inter-quartile range (≤ 2) for section 4. As part of this process, voting permitted the Core Group to later make minor clarifications to the model (Stage 6). A final model was presented to the entire panel, with an additional open comment period of two weeks ensuring representation of emerged categories.

In Stage 7, four authors (MSK, MD, OA, LR) established specific criteria to define each HOT ^(29)^, as seen in Supplement 5. These criteria defined what explicitly and implicitly constituted each higher-order category, what explicitly was *not* part of each category. A new form/template was then created to re-screen the searchable pdf of all manuscripts for tasks and their properties (e.g., program delivery mode: in-person, home line; format: 1-on-1, group) in each category. The re-review and recode using these forms were completed by two authors (MSK, OA). In some cases, emails were sent to corresponding authors for clarification on study details.

**Supplement 5.** Criteria for task identification and classification.

| Supplement 5. Criteria for task identification and classification. |  |  |  |  |  |
| --- | --- | --- | --- | --- | --- |
|  | Task Category from Table 2 | Does NOT include | Explicit (X symbol) | Implicit ( / symbol) suggesting strongly that it was conducted | Notes and search criteria |
| 1 | Risk Factor and needs identification | Does **not** include:<br>Limitations due to age, weight status and various eligibility factors.<br>Signed approval from PCP/surgeon for exercise | 1. Exercise health & musculoskeletal screening,<br>2. determination of contraindications<br>3. Needs identification<br>Includes:<br>PAR-Q, | Inclusion/ exclusion criteria suggestive that a clinical screening was conducted. | Search for words: RISK / SCREENING / NEEDS / CONTRAINDICATIONS<br>In some studies, no additional exercise-related health screening was conducted (e.g., Parikh; Rothwell) |
| 2 | Exercise-related health assessment | Does **not** include:<br>Functional testing, like 6-minute walk tests | physical by a sports physician<br>Vitals (Blood pressure) – e.g., Funderburk<br>Bruce protocols<br>Modified-Bruce protocols | VO2max data or similar was reported |  |
| 3 | Body Composition Assessment | Does **not** include:<br>Does not include simple weight, height and BMI. | Tests of percent body fat and resting metabolic rate and waist and hip circumferences. | Data suggestive of test completion |  |
| 4 | Fitness testing | Does **not** include:<br>“Perceived” fitness | Other non-graded exercise tests, like the 6MWT or the Cooper Test. | Data suggestive of test completion |  |
| 5 | Lifestyle physical activity and sedentary behaviors assessment | Does **not** include:<br>Gym attendance | Accelerometry (i.e., Actigraph);<br>Fitness trackers (i.e., Fitbits)<br>Pedometry<br>Diaries / logs of activity | Step data included | Search for words: STEPS |
| 6 | Fitness and Health Education | Does **not** include:<br>Information about the program | Structured exercise curriculum | Evidence of curriculum (e.g., table of contents) |  |
|  |  |  | Includes “exercise guidelines” as provided in a control group for a RCT.<br>Exercise instructions for home. |  |  |
| 7 | Physical Activity Instruction | Does **not** include:<br>Structured exercise program | Includes:<br>Familiarization<br>Dumbbells, resistance bands, bands, | List of exercises that would infer that patient would need instruction |  |
| 8 | Exercise Prescription | Does **not** include:<br>Outside physical activity that a patient would routinely conduct on their own.<br>Exercise counseling about exercise<br>Exercise or PA education | Any exercise outside of the structured/supervised training program, as provided by the exercise professional.<br>Includes access to facility during non-training times | Text highly suggestive that a home program was being conducted | Search for words:<br>HOME, OUTSIDE |
|  |  |  | Prescribed exercise training program defined by exercise variables, such as:<br>Minutes/time (duration)<br>Frequency of exercise<br>Type/Mode<br>Etc.<br>Early ambulation program | Text highly suggestive that a standardized program was being conducted. |  |
| 9 | Personalization | Does **not** include:<br>General guidelines are given to all/most patients<br>Different guidelines/instructions for two or more different groups | Customization, individualization and/or modification based on needs/ health status<br>Modifications based on pain and tolerance<br>Implied if they use heart rate % to prescribe the exercise.<br>“Optimization” of exercise program | Heart rate zone provision<br>If a case study, it is assumed to be individualized for the patient. | Search for words:<br>Individualization, Modification, Customization |
|  |  | Does **not** include:<br>Standardized exercise prescription that does not change over time. | Weekly, bi-weekly or monthly changes based on a variety of factors, like improvements in fitness or less limitations. | Adjustments made but the criteria by which the adjustments are made is not clear, or it is not applicable for all patients (only some were progressed). | Search for words:<br>Progression<br>Increment* |
| 10 | Metabolic Assessment | Does **not** include:<br>HR <u>prescription</u> only | Also includes BP and SPO2 and glucose | Data suggestive | Search for words:<br>Heart rate |
|  |  |  | Additional action if parameters are out-of-expected range. |  | Glucose<br>Blood sugar |
| 11 | Monitoring | Does **not** include:<br>Any activities in which patients were not within sight. | Includes orientation and familiarization and training to ensure safety and low abdominal pressure<br>Supervision / Supervised exercise training, including instruction, guiding and feedback<br>Teaching the Borg exertion scale to self-monitor | Text highly suggestive that feedback was being provided. | In many studies, it is not clear if some monitoring was passive or strictly active (i.e., passive monitoring from a desk/station at a gym vs. continuous supervision of each patient. |
| 12 | Counseling Technique | Does **not** include:<br>Education<br>Passing out guidelines<br>General advice | Instructions on goal setting, motivational interviewing or other techniques<br>Contingency planning ("if this happens, do this...")<br>Strategies to build confidence through experiences | Evidence of strategies (e.g., from a table) |  |
| 13 | Psychosocial Support | Does **not** include:<br>Therapy<br>Other services necessitating psychiatry licenses | Onward Referral<br>Support of Psycho/Social Worker | Tables with psychological data in it in methods or results, information in regards to psychosocial assessments |  |
| 14 | Dietary Support | Does **not** include:<br>Therapy<br>Other treatments necessitating licensing of dieticians | Education of basic consulting on bariatric dietary basics per-post surgery, (PPO, Supplements, Macros, Importance of vitamins);<br>energy balance<br>Assessment<br>Onward referral | Lists of information about macronutrients, etc. |  |

## RESULTS

A total of 63 studies were found describing exercise programs in MBS, representing 50 unique interventions (many studies had more than one published report). A further search to include all articles related to these interventions increased the total to 71 with 57% published after 2015. Over half of the studies were from just two countries – the USA (39%) and Brazil (14%). Exercise programs were mainly conducted in teaching hospitals (38%). The majority of articles were “proof of concept” (i.e., addressing the question of whether exercise is beneficial for weight loss) as opposed to implementation. The main component of these programs was supervised exercise training (65%). Later article additions increased this total to 83, which were analyzed later. See the Tables and reference list in Supplement 6.

## Supplement 6 with citations (see below)

**Table 1A.**
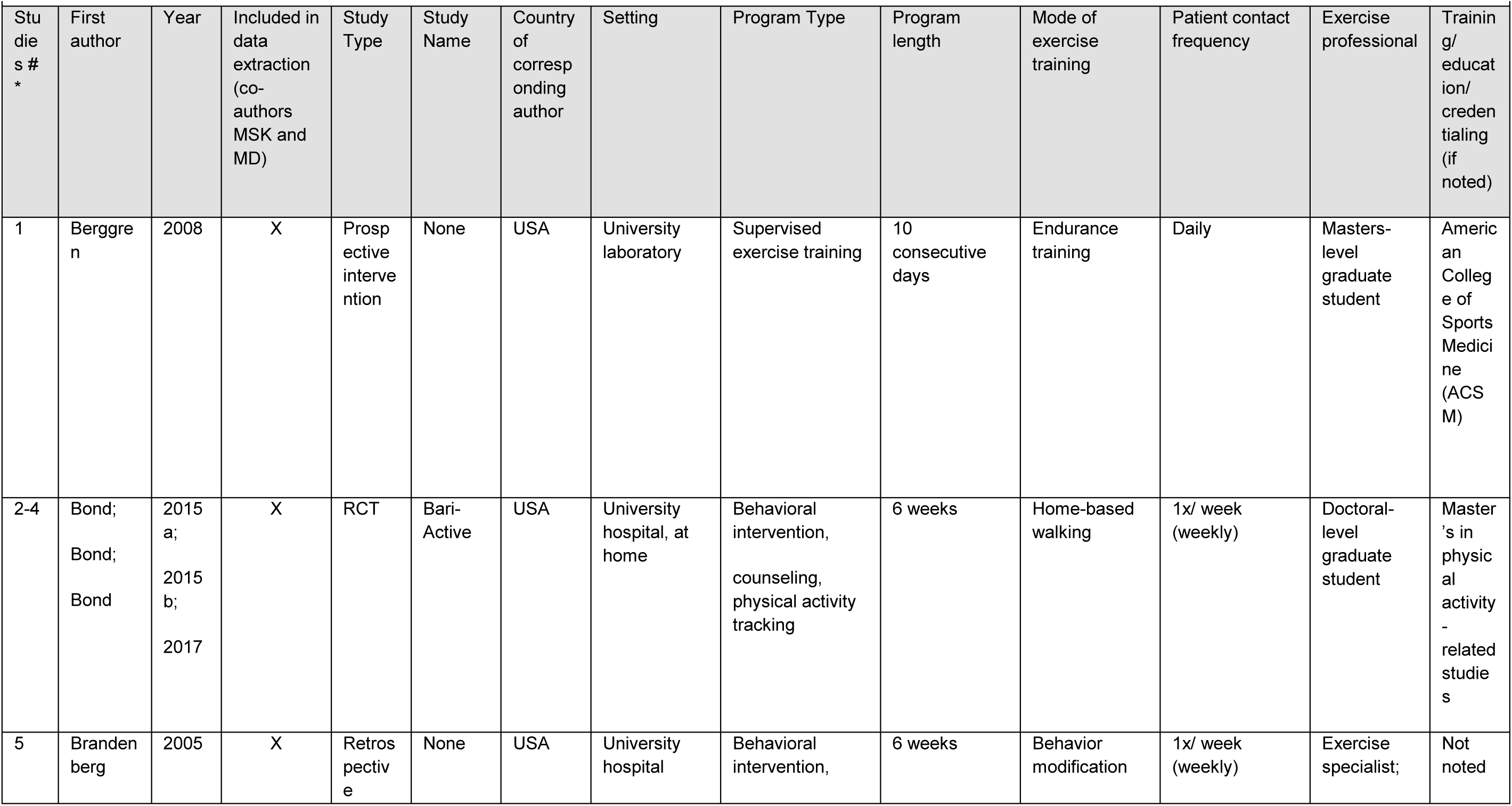
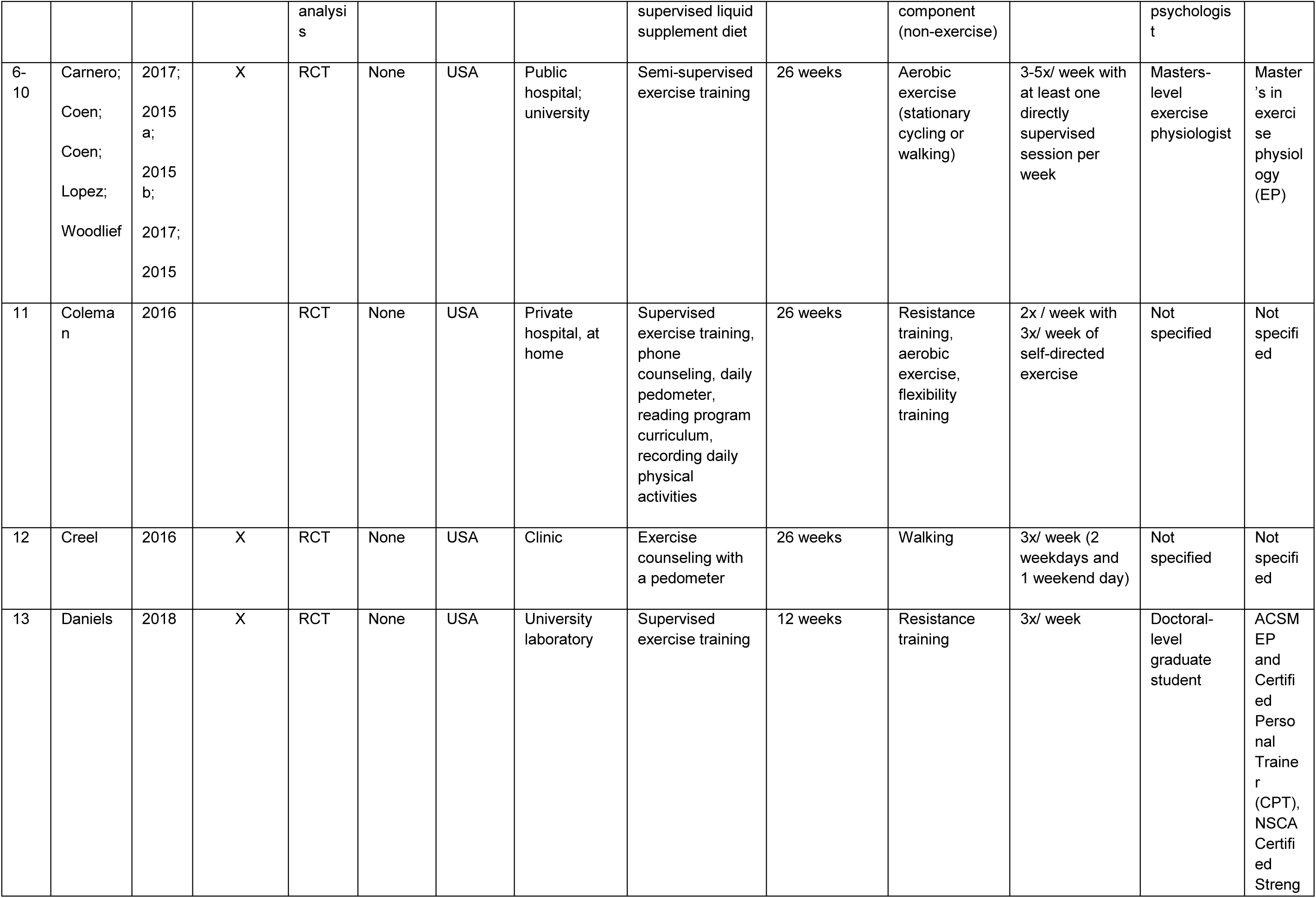
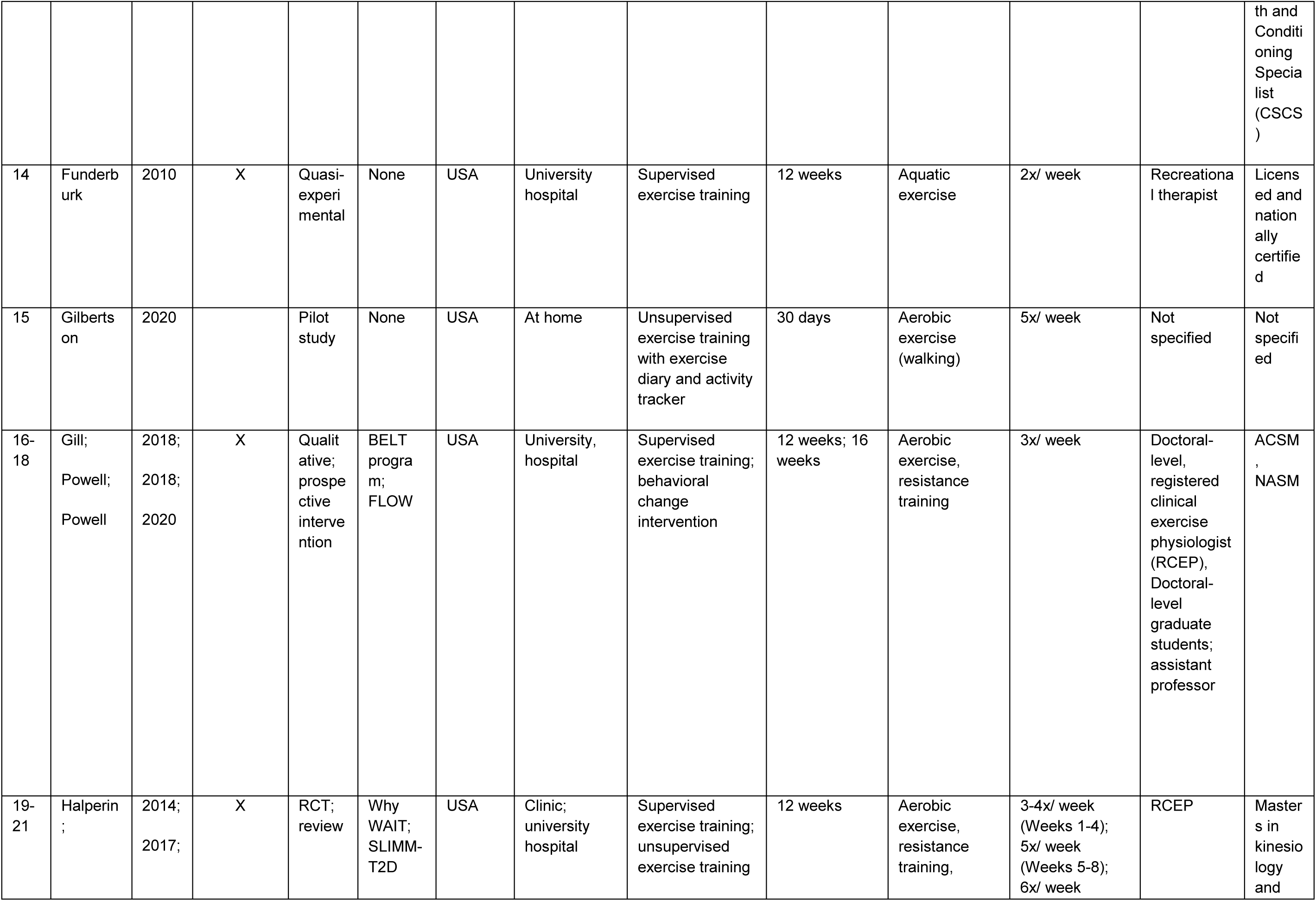
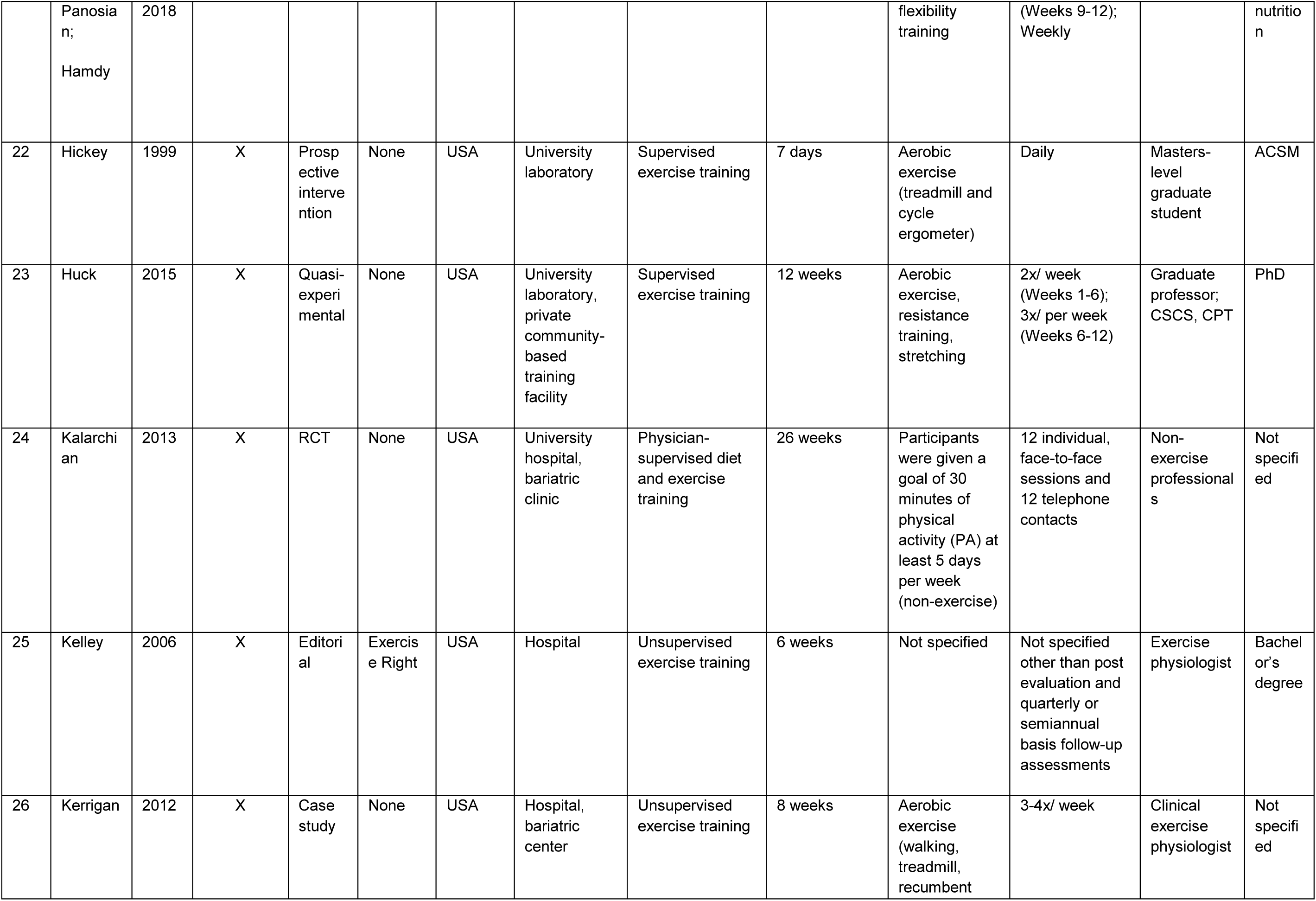
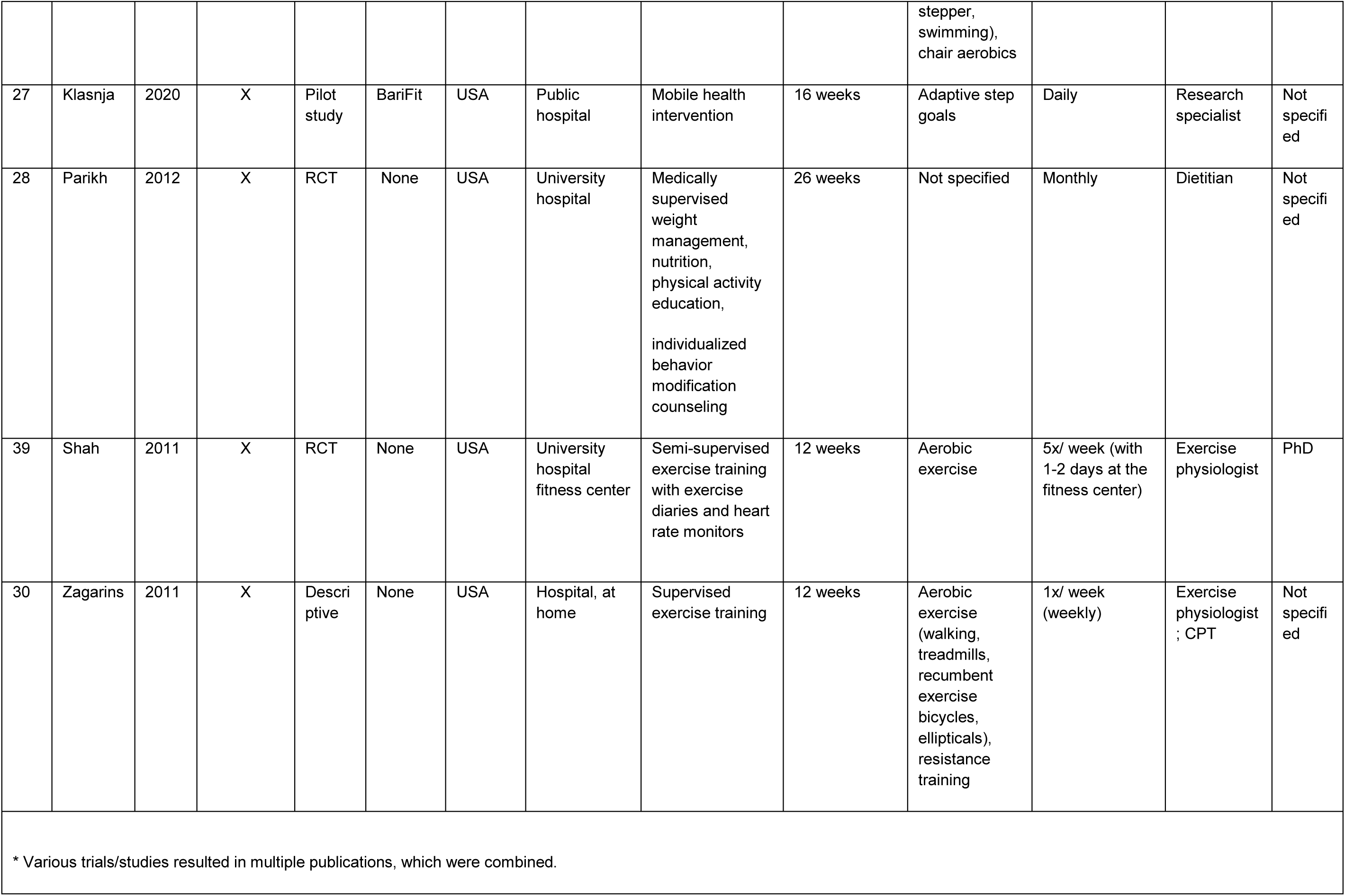
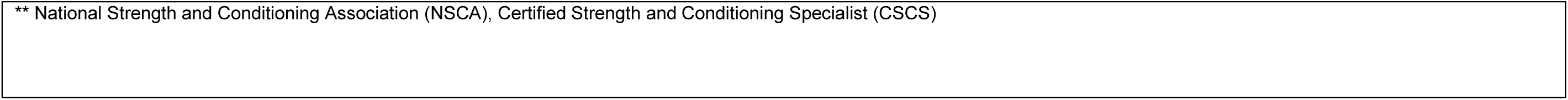
Details of studies from the USA.

| Table 1A. Details of studies from the USA |  |  |  |  |  |  |  |  |  |  |  |  |  |
| --- | --- | --- | --- | --- | --- | --- | --- | --- | --- | --- | --- | --- | --- |
| Studies # | First author | Year | Included in data extraction (co-authors MSK and MD) | Study Type | Study Name | Country of corresponding author | Setting | Program Type | Program length | Mode of exercise training | Patient contact frequency | Exercise professional | Training/education/credentialing (if noted) |
| 1 | Berggren | 2008 | X | Prospective intervention | None | USA | University laboratory | Supervised exercise training | 10 consecutive days | Endurance training | Daily | Masters-level graduate student | American College of Sports Medicine (ACSM) |
| 2-4 | Bond;<br>Bond;<br>Bond | 2015 a;<br>2015 b;<br>2017 | X | RCT | Bari-Active | USA | University hospital, at home | Behavioral intervention, counseling, physical activity tracking | 6 weeks | Home-based walking | 1x/ week (weekly) | Doctoral-level graduate student | Master's in physical activity-related studies |
| 5 | Brandenberg | 2005 | X | Retrospective | None | USA | University hospital | Behavioral intervention, | 6 weeks | Behavior modification | 1x/ week (weekly) | Exercise specialist; | Not noted |

|  |  |  |  | analysis |  |  |  | supervised liquid supplement diet |  | component (non-exercise) |  | psychologist |  |
| --- | --- | --- | --- | --- | --- | --- | --- | --- | --- | --- | --- | --- | --- |
| 6-10 | Carnero; Coen; Coen; Lopez; Woodlief | 2017; 2015 a; 2015 b; 2017; 2015 | X | RCT | None | USA | Public hospital; university | Semi-supervised exercise training | 26 weeks | Aerobic exercise (stationary cycling or walking) | 3-5x/ week with at least one directly supervised session per week | Masters-level exercise physiologist | Master's in exercise physiology (EP) |
| 11 | Coleman | 2016 |  | RCT | None | USA | Private hospital, at home | Supervised exercise training, phone counseling, daily pedometer, reading program curriculum, recording daily physical activities | 26 weeks | Resistance training, aerobic exercise, flexibility training | 2x / week with 3x/ week of self-directed exercise | Not specified | Not specified |
| 12 | Creel | 2016 | X | RCT | None | USA | Clinic | Exercise counseling with a pedometer | 26 weeks | Walking | 3x/ week (2 weekdays and 1 weekend day) | Not specified | Not specified |
| 13 | Daniels | 2018 | X | RCT | None | USA | University laboratory | Supervised exercise training | 12 weeks | Resistance training | 3x/ week | Doctoral-level graduate student | ACSM EP and Certified Personal Trainer (CPT), NSCA Certified Strength th and<br>Condi<br>oning<br>Specia<br>list<br>(CSCS<br>) |
| 14 | Funderb<br>urk | 2010 | X | Quasi-<br>experi<br>mental | None | USA | University<br>hospital | Supervised<br>exercise training | 12 weeks | Aquatic<br>exercise | 2x/ week | Recreational<br>therapist | Licens<br>ed and<br>nation<br>ally<br>certifie<br>d |
| 15 | Gilberts<br>on | 2020 |  | Pilot<br>study | None | USA | At home | Unsupervised<br>exercise training<br>with exercise<br>diary and activity<br>tracker | 30 days | Aerobic<br>exercise<br>(walking) | 5x/ week | Not<br>specified | Not<br>specifi<br>ed |
| 16-18 | Gill;<br>Powell;<br>Powell | 2018;<br>2018;<br>2020 | X | Qualit<br>ative;<br>prospe<br>ctive<br>interve<br>ntion | BELT<br>progra<br>m;<br>FLOW | USA | University,<br>hospital | Supervised<br>exercise training;<br>behavioral<br>change<br>intervention | 12 weeks; 16<br>weeks | Aerobic<br>exercise,<br>resistance<br>training | 3x/ week | Doctoral-level,<br>registered<br>clinical<br>exercise<br>physiologist<br>(RCEP),<br>Doctoral-level<br>graduate<br>students;<br>assistant<br>professor | ACSM<br>,<br>NASM |
| 19-21 | Halperin<br>; | 2014;<br>2017; | X | RCT;<br>review | Why<br>WAIT;<br>SLIMM-T2D | USA | Clinic;<br>university<br>hospital | Supervised<br>exercise training;<br>unsupervised<br>exercise training | 12 weeks | Aerobic<br>exercise,<br>resistance<br>training, | 3-4x/ week<br>(Weeks 1-4);<br>5x/ week<br>(Weeks 5-8);<br>6x/ week | RCEP | Master<br>s in<br>kinesio<br>logy<br>and |
|  | Panosia<br>n;<br>Hamdy | 2018 |  |  |  |  |  |  |  | flexibility<br>training | (Weeks 9-12);<br>Weekly |  | nutritio<br>n |
| 22 | Hickey | 1999 | X | Prosp<br>ective<br>interve<br>nction | None | USA | University<br>laboratory | Supervised<br>exercise training | 7 days | Aerobic<br>exercise<br>(treadmill and<br>cycle<br>ergometer) | Daily | Masters-level<br>graduate<br>student | ACSM |
| 23 | Huck | 2015 | X | Quasi-<br>experi<br>mental | None | USA | University<br>laboratory,<br>private<br>community-<br>based<br>training<br>facility | Supervised<br>exercise training | 12 weeks | Aerobic<br>exercise,<br>resistance<br>training,<br>stretching | 2x/ week<br>(Weeks 1-6);<br>3x/ per week<br>(Weeks 6-12) | Graduate<br>professor;<br>CSCS, CPT | PhD |
| 24 | Kalarchi<br>an | 2013 | X | RCT | None | USA | University<br>hospital,<br>bariatric<br>clinic | Physician-<br>supervised diet<br>and exercise<br>training | 26 weeks | Participants<br>were given a<br>goal of 30<br>minutes of<br>physical<br>activity (PA) at<br>least 5 days<br>per week<br>(non-exercise) | 12 individual,<br>face-to-face<br>sessions and<br>12 telephone<br>contacts | Non-exercise<br>professional<br>s | Not<br>specifi<br>ed |
| 25 | Kelley | 2006 | X | Editori<br>al | Exercis<br>e Right | USA | Hospital | Unsupervised<br>exercise training | 6 weeks | Not specified | Not specified<br>other than post<br>evaluation and<br>quarterly or<br>semiannual<br>basis follow-up<br>assessments | Exercise<br>physiologist | Bachel<br>or's<br>degree |
| 26 | Kerrigan | 2012 | X | Case<br>study | None | USA | Hospital,<br>bariatric<br>center | Unsupervised<br>exercise training | 8 weeks | Aerobic<br>exercise<br>(walking,<br>treadmill,<br>recumbent stepper, swimming), chair aerobics | 3-4x/ week | Clinical<br>exercise<br>physiologist | Not<br>specifi<br>ed |
| 27 | Klasnja | 2020 | X | Pilot study | BariFit | USA | Public hospital | Mobile health intervention | 16 weeks | Adaptive step goals | Daily | Research specialist | Not specified |
| 28 | Parikh | 2012 | X | RCT | None | USA | University hospital | Medically supervised weight management, nutrition, physical activity education, individualized behavior modification counseling | 26 weeks | Not specified | Monthly | Dietitian | Not specified |
| 39 | Shah | 2011 | X | RCT | None | USA | University hospital fitness center | Semi-supervised exercise training with exercise diaries and heart rate monitors | 12 weeks | Aerobic exercise | 5x/ week (with 1-2 days at the fitness center) | Exercise physiologist | PhD |
| 30 | Zagarins | 2011 | X | Descriptive | None | USA | Hospital, at home | Supervised exercise training | 12 weeks | Aerobic exercise (walking, treadmills, recumbent exercise bicycles, ellipticals), resistance training | 1x/ week (weekly) | Exercise physiologist ; CPT | Not specified |
| * Various trials/studies resulted in multiple publications, which were combined. |  |  |  |  |  |  |  |  |  |  |  |  |  |

\*\* National Strength and Conditioning Association (NSCA), Certified Strength and Conditioning Specialist (CSCS)

**Table 1B.**
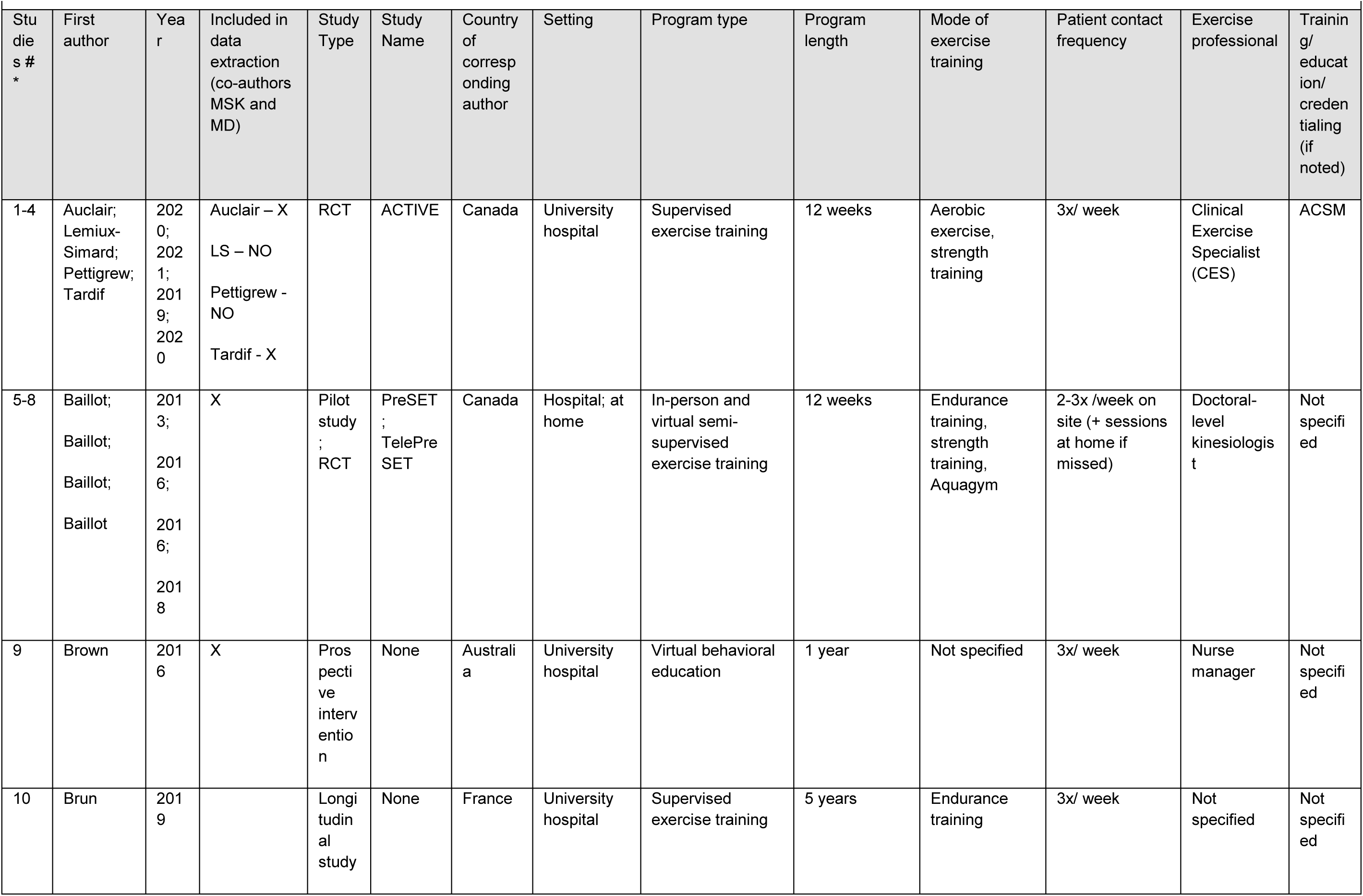
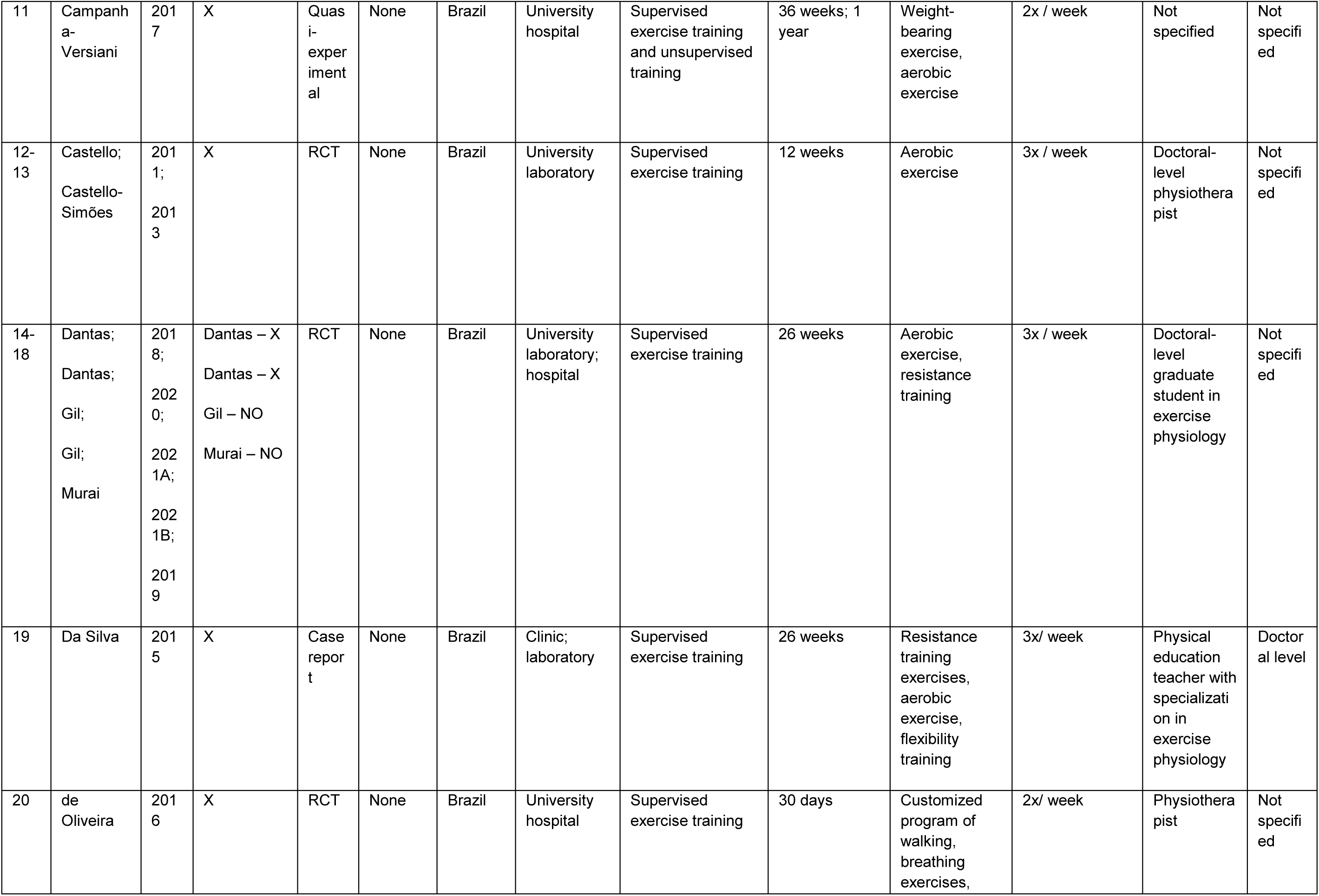
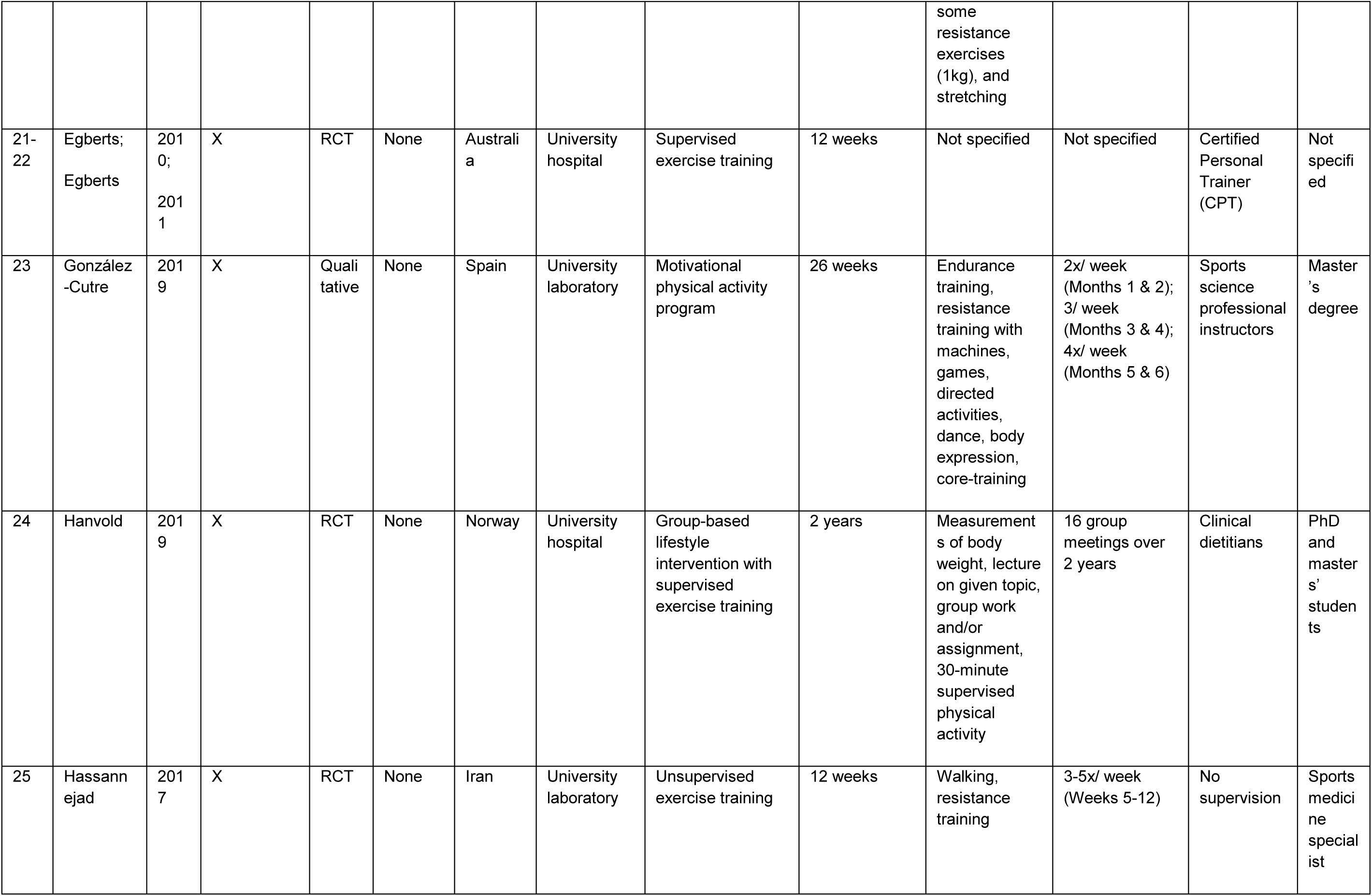
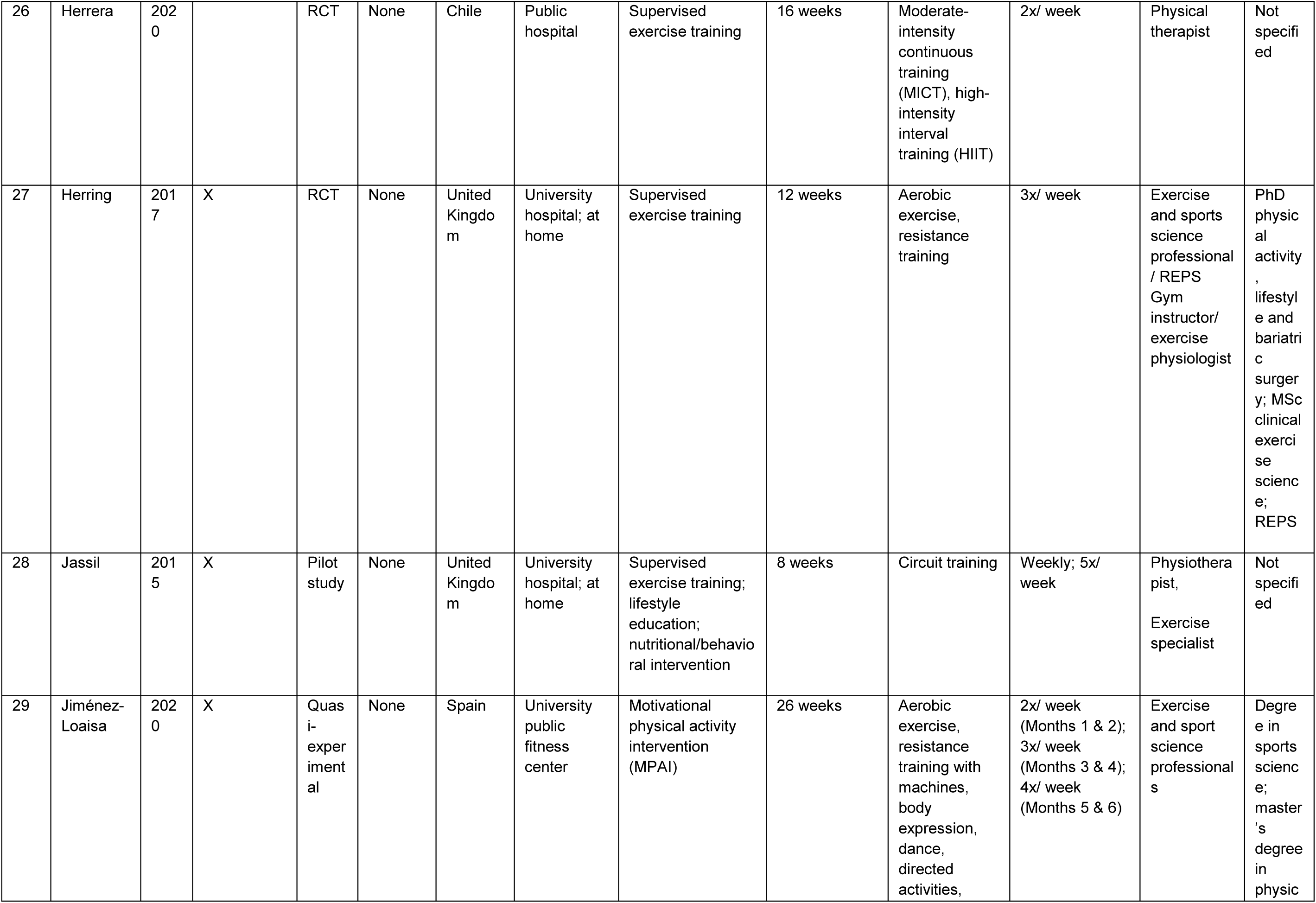
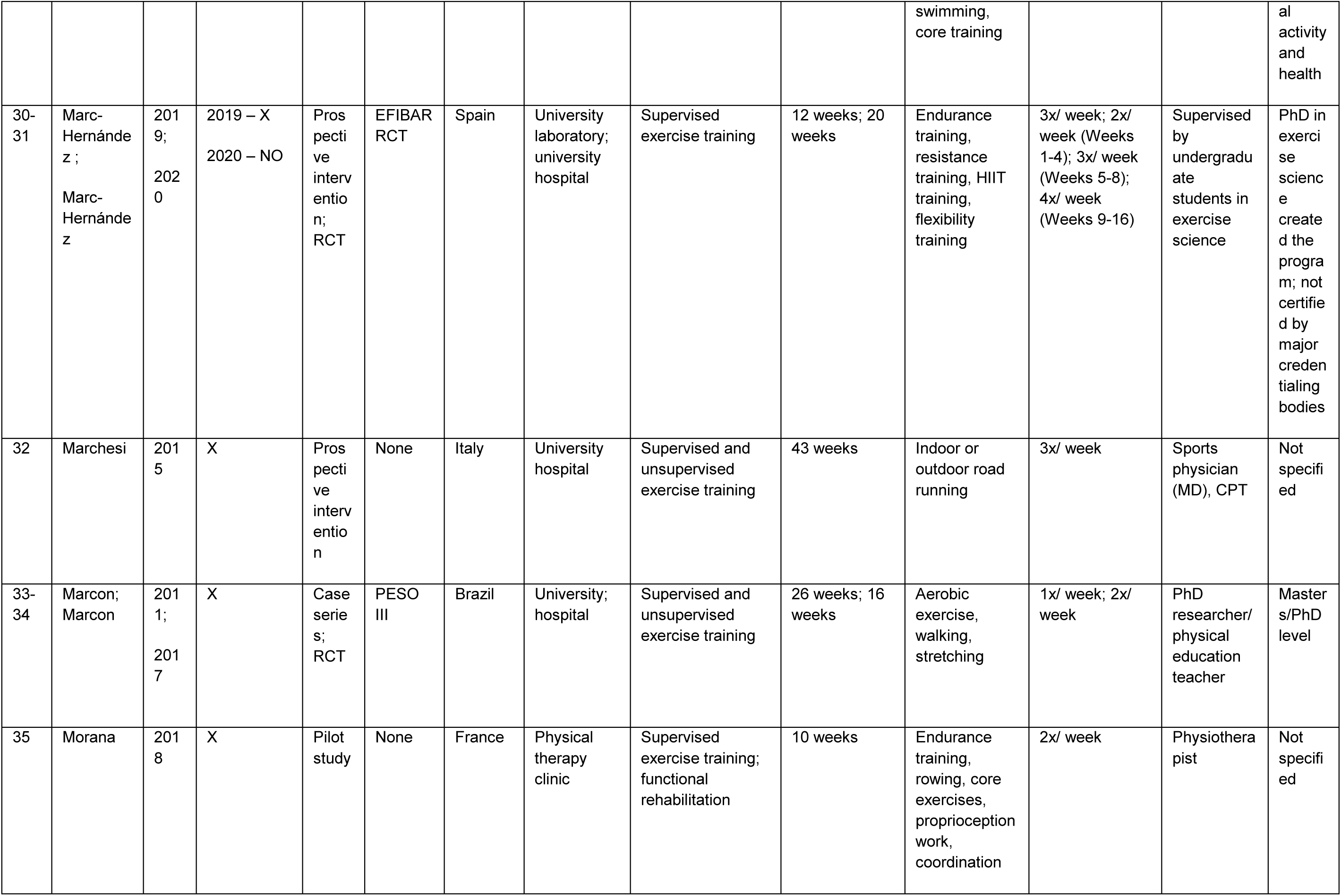
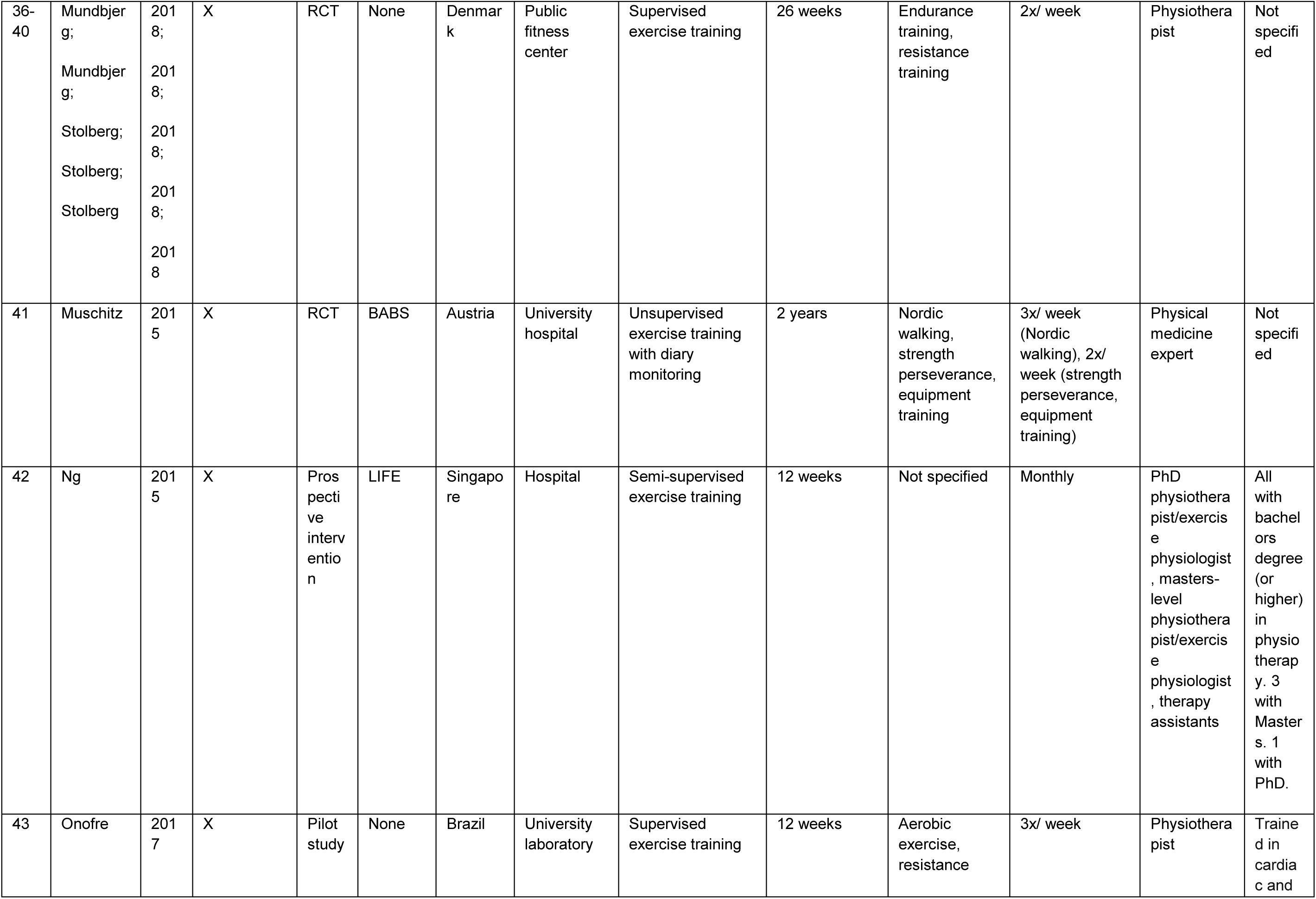
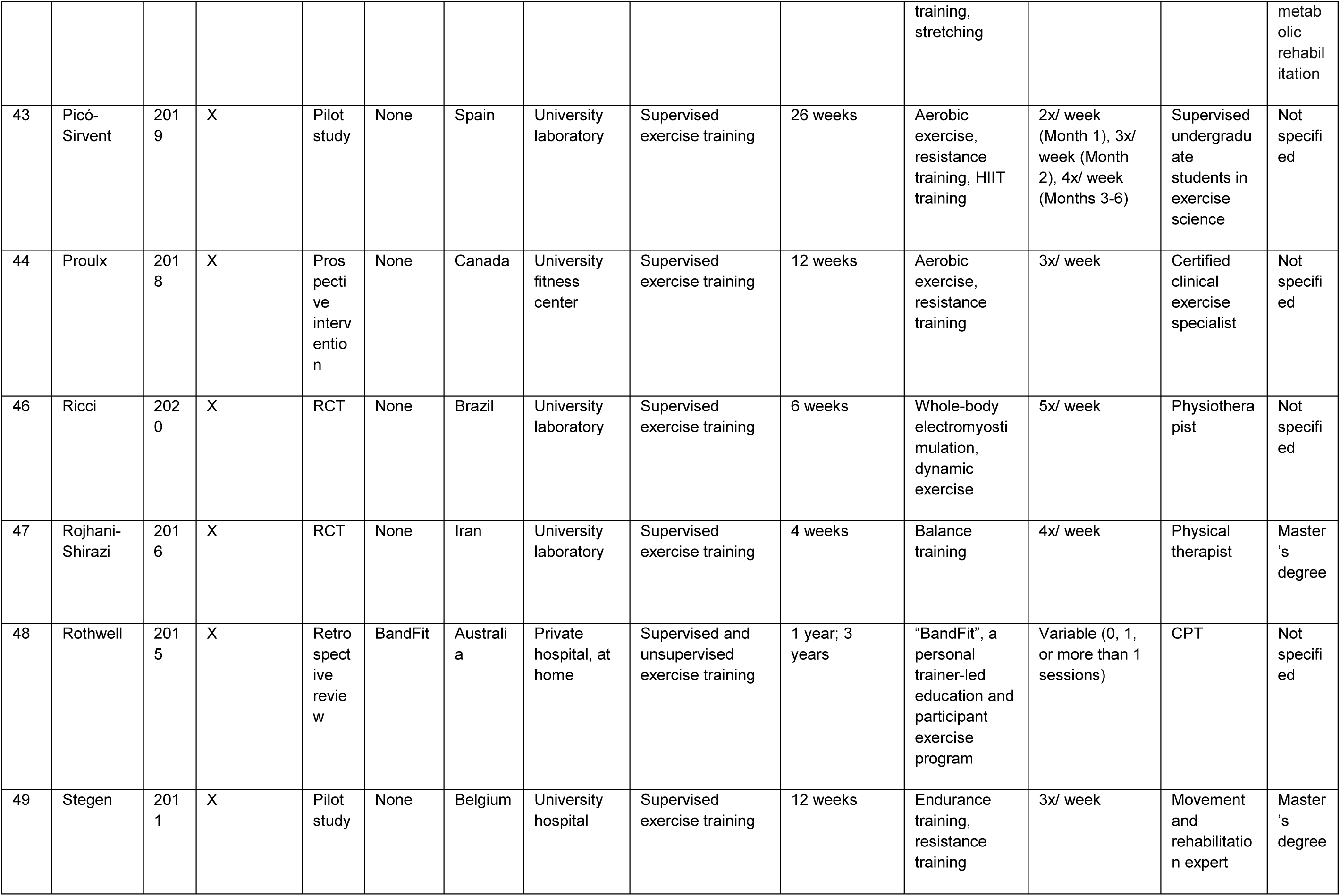
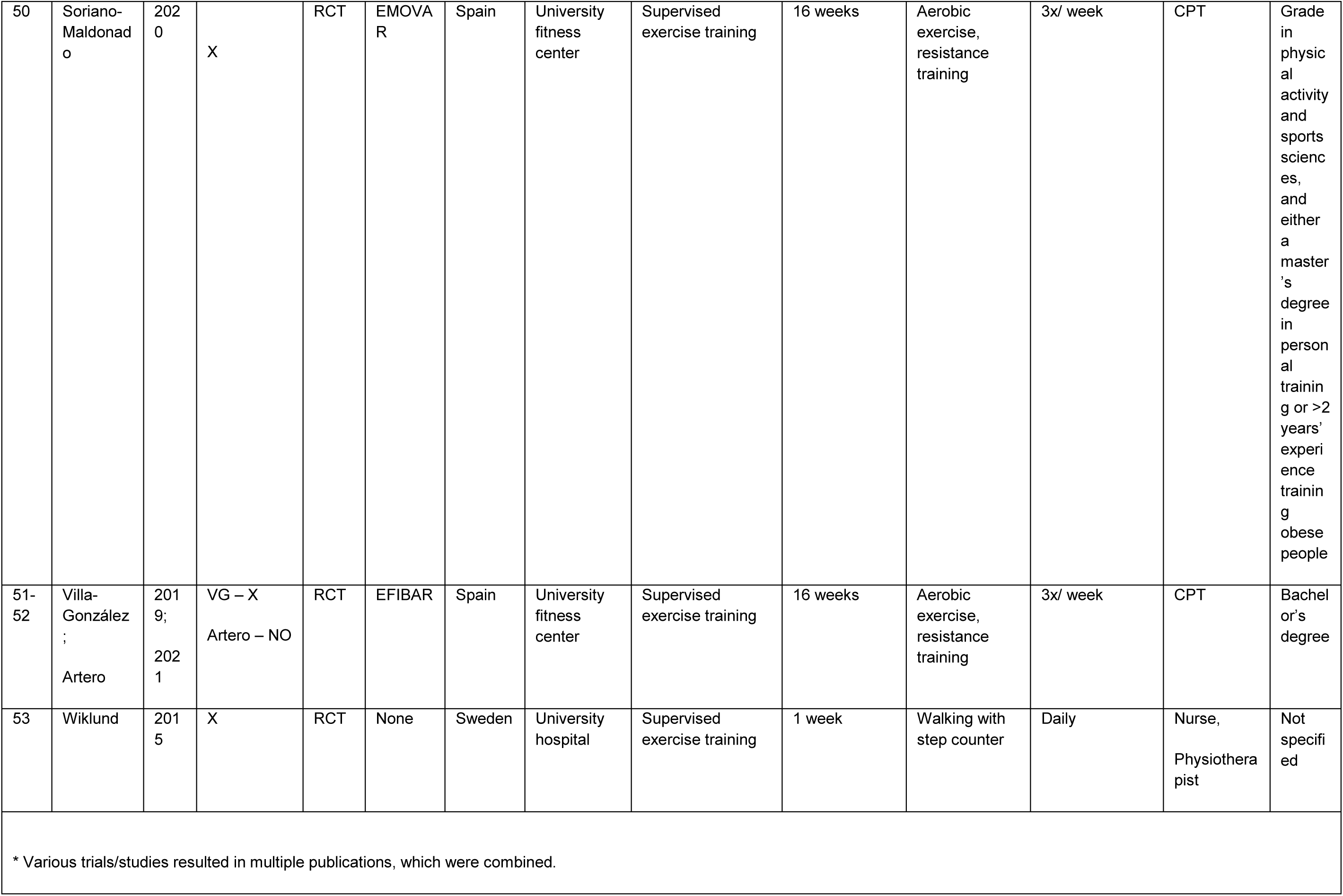
Details of studies from outside the USA.

## Classification and qualifications of exercise professionals

The exercise professional delivering the PA programming was firstly an exercise physiologist in the USA (28%; person with graduate degree in EP and /or certification) and a physiotherapist/physical therapist in the rest of the world (43%). A number of other exercise professionals (e.g., physical educators, coaches, personal trainers) were also involved. Qualifications and credentials were difficult to ascertain due to underreporting. However, it was discovered (through details in studies and/or email inquiries) that 6 studies used bachelor’s-level exercise professionals, 9 used Masters-level, 9 used PhD, 2 used medical doctors (MD). In seven cases, exercise physiologists were specifically credentialed by the American College of Sports Medicine (ACSM) and in one case by the National Strength and Conditioning Association (NSCA).

## Task data extraction

Data extraction resulted in 385 tasks across all studies, 43 of which were unique. The 5 most common tasks were: real-time supervision of exercise (36 studies), fitness testing (31), [exercise] prescription (27), heart rate monitoring (21), and physical activity counseling (21). Less commonly described in the literature were: exercise prescription for home, exercise behavioral counseling, and metabolic testing.

## Content analysis / Card sort

Respondents sorted tasks into an average of 11.8 bins, each bin had an average of 3.6 items, and there was an average of 1.8 items deleted per respondent. Respondents suggested 34 items to add (3.4/respondent). Unique items were considered in later rounds. See Supplement 7.

**Supplement 7.** Summary of bin sort per analyst.

| **Supplement 7. Summary of bin sort per analyst.** |  |  |  |  |  |  |  |  |  |  |  |
| --- | --- | --- | --- | --- | --- | --- | --- | --- | --- | --- | --- |
| **Rater #** | **1** | **2** | **3** | **4** | **5** | **6** | **7** | **8** | **9** | **10** | **Average** |
| **Analyst initials** | **DB** | **CG** | **EB** | **BG** | **MSK** | **LR** | **JH** | **LH** | **RM** | **MD** |  |
| **Bins completed (#)** | 12 | 14 | 10 | 11 | 14 | 11 | 10 | 11 | 10 | 15 | 11.8 |
| **Bins w/ 8 items** | 0 | 1 | 1 | 0 | 0 | 0 | 1 | 0 | 0 | 0 | 0.3 |
| **Bins w/ 7 items** | 0 | 1 | 1 | 0 | 0 | 0 | 1 | 0 | 0 | 0 | 0.3 |
| **Bins w/ 6 items** | 0 | 0 | 0 | 1 | 0 | 3 | 1 | 3 | 2 | 1 | 1.1 |
| **Bins w/ 5 items** | 5 | 0 | 1 | 2 | 1 | 0 | 1 | 2 | 3 | 1 | 1.6 |
| **Bins w/ 4 items** | 1 | 1 | 2 | 2 | 2 | 2 | 1 | 0 | 2 | 4 | 1.7 |
| **Bins w/ 3 items** | 3 | 5 | 3 | 1 | 5 | 4 | 2 | 2 | 2 | 1 | 2.8 |
| **Bins w/ 2 items** | 1 | 3 | 2 | 2 | 4 | 0 | 1 | 3 | 1 | 5 | 2.2 |
| **Bins w/ 1 items** | 2 | 3 | 0 | 3 | 1 | 2 | 2 | 1 | 0 | 3 | 1.7 |
| **Items to discard** | 0 | 0 | 2 | 7 | 3 | 2 | 2 | 2 | 0 | 0 | 1.8 |
| **Ave items/bin\*** | 3.5 | 3.1 | 4.1 | 3.3 | 2.9 | 3.7 | 4.1 | 3.7 | 4.3 | 2.9 | 3.6 |
| **Items missing to add** | 0 | 14 | 0 | 7 | 0 | 5 | 6 | 0 | 2 | 0 | 3.4 |
| * ([# items – discarded items]/bins completed) |  |  |  |  |  |  |  |  |  |  |  |

A heat map of the associations of the original 43 exercise professional job tasks is provided in Supplement 8. The items with the strongest association (9 out of 10 raters agreed) were strength testing and physical function testing. Eight out of 10 raters indicated that the following 3 tasks were paired: strength testing/balance testing, fitness testing/function testing, prescription for home/ prescription-individual. Seven raters agreed on 5 pairs, 6 raters - 10 pairs, 5 raters −17 pairs, etc.

**Supplement 8.**
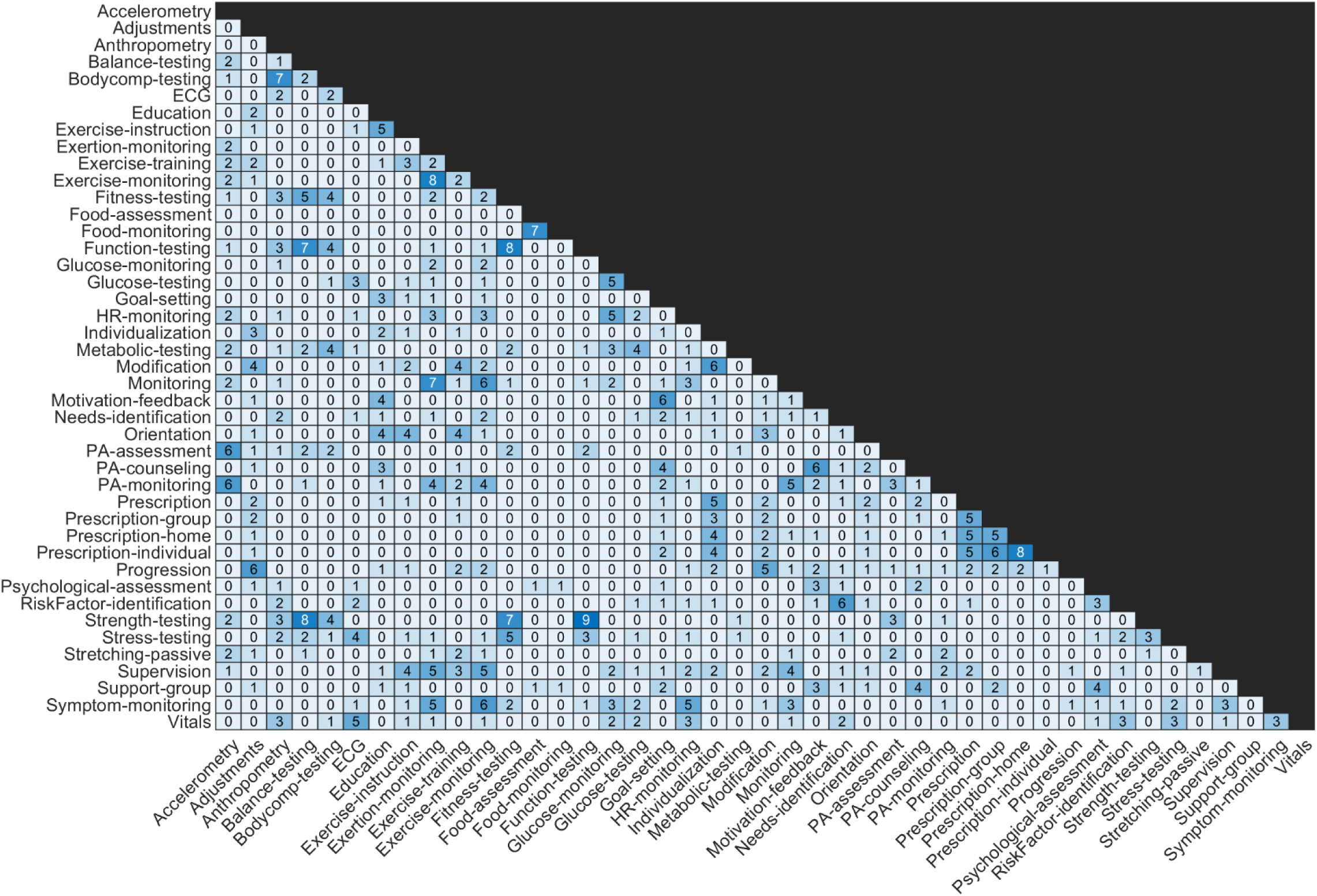
Heat map demonstrating task associations within sorting bins.

The hierarchical cluster analysis based on this matrix generated two models, which are shown in Supplement 9. Model B had a better fit index than Model A, as quantified by the cophenetic correlation coefficient (c = 0.85 and 0.50, respectively), a common statistic that measures the faithfulness with which the similarities between individual items in the original matrix were represented visually in the final dendrogram.

**Supplement 9.**
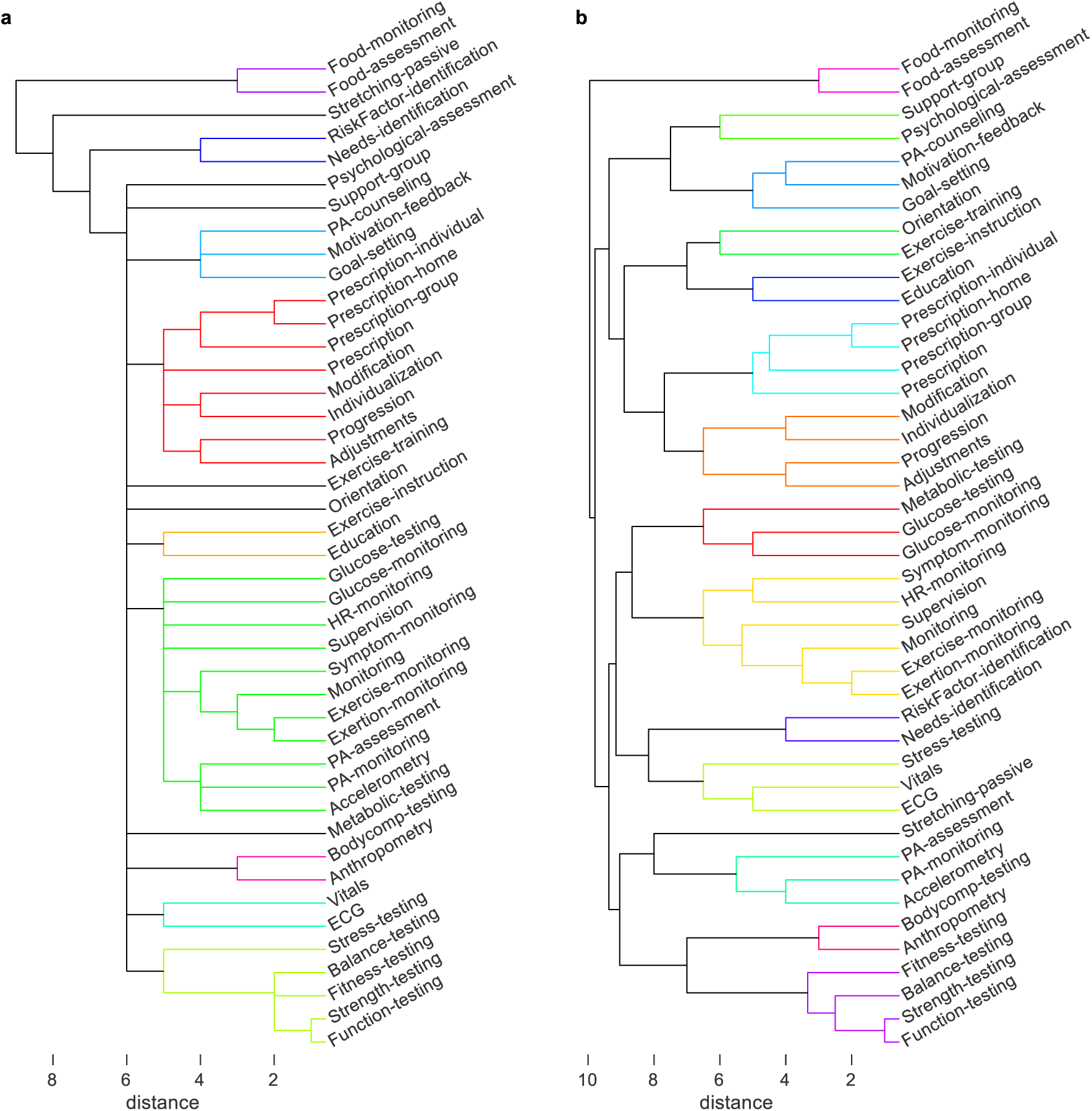
Alternative models (A and B) generated by the hierarchical cluster analysis and deliberated in the Delphi process.

## Delphi process / Core Group

Respondents approved Model B. They indicated that “Passive stretching” should be eliminated and “exercise prescription adherence” should be included. The Core Group eliminated the general “prescription” task. Therefore, the final model included 42 patient-centered exercise tasks (i.e., lower order themes), which divided into 14 higher order themes and 7 SHOTs. The final SHOTs were: (1) Exercise-related health assessment, (2) Body composition and physical fitness assessment, (3) Lifestyle physical activity and sedentary behavior assessment, (4) Education, instruction, and prescription, (5) Exercise monitoring, (6) Behavioral counseling and psychosocial support, (7) Dietary support. The final model passed with a unanimous final vote. See Table 1, Figure 2, and Supplements 10-11 for the final model, with descriptions of the SHOTs. Task categories were not ranked, per group consensus, but were further sorted by temporal precedence in a clinical setting (starting with basic assessments).

**Figure 2.**
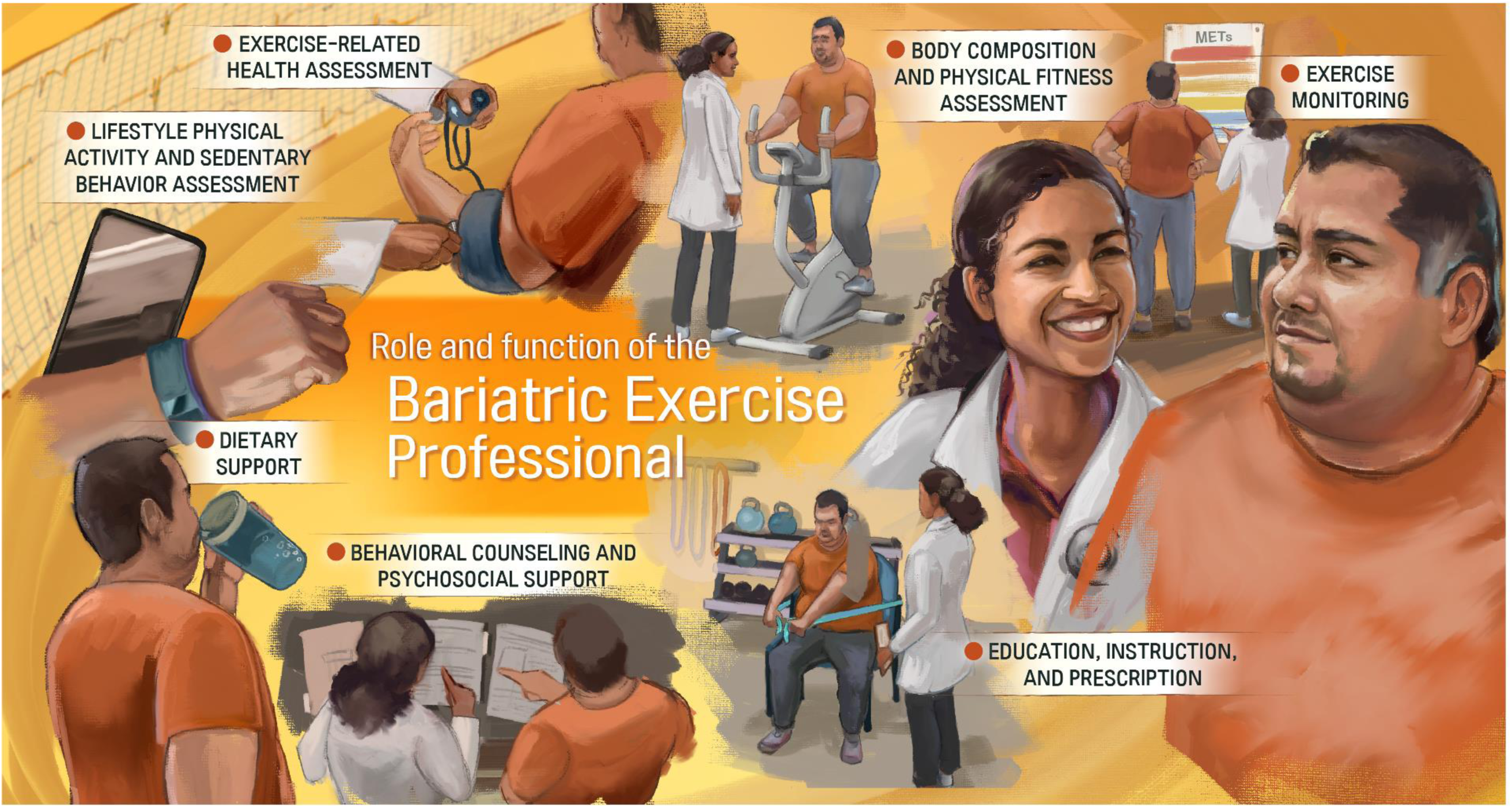
Illustrated final model (Version B) for clinical use. Note: Certain aspects of tasks may be outside the scope of training for some specific exercise professions, and patients with multiple comorbidities would be better served seeing a clinical exercise physiologist as opposed to a personal trainer.

**Table 1.**
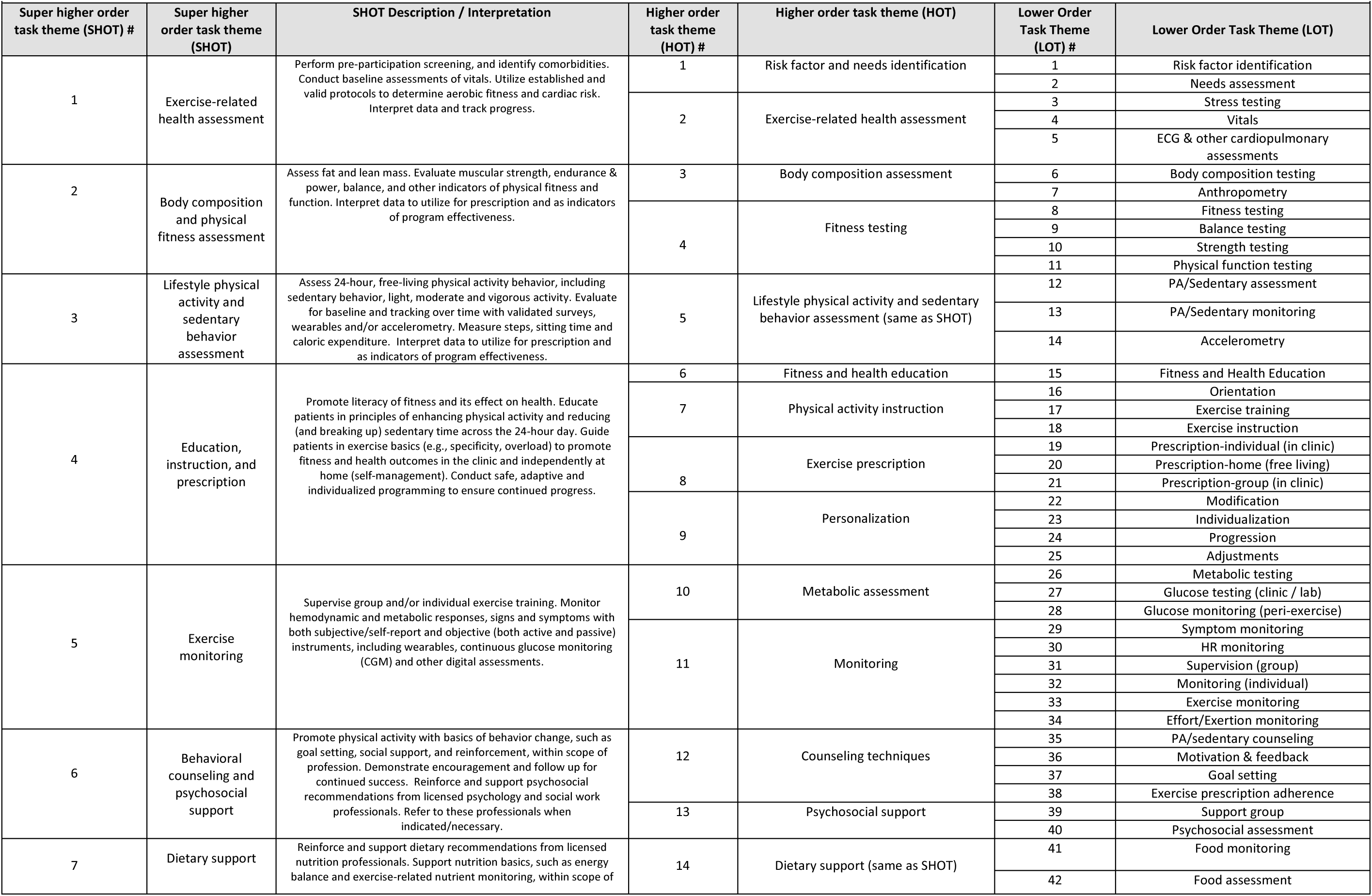
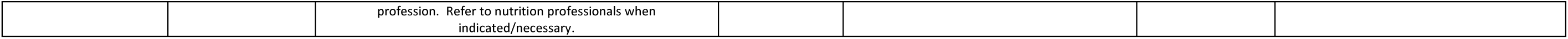
Final model with descriptions of the Super Higher Order Task (SHOT) themes.

**Supplement 10.**
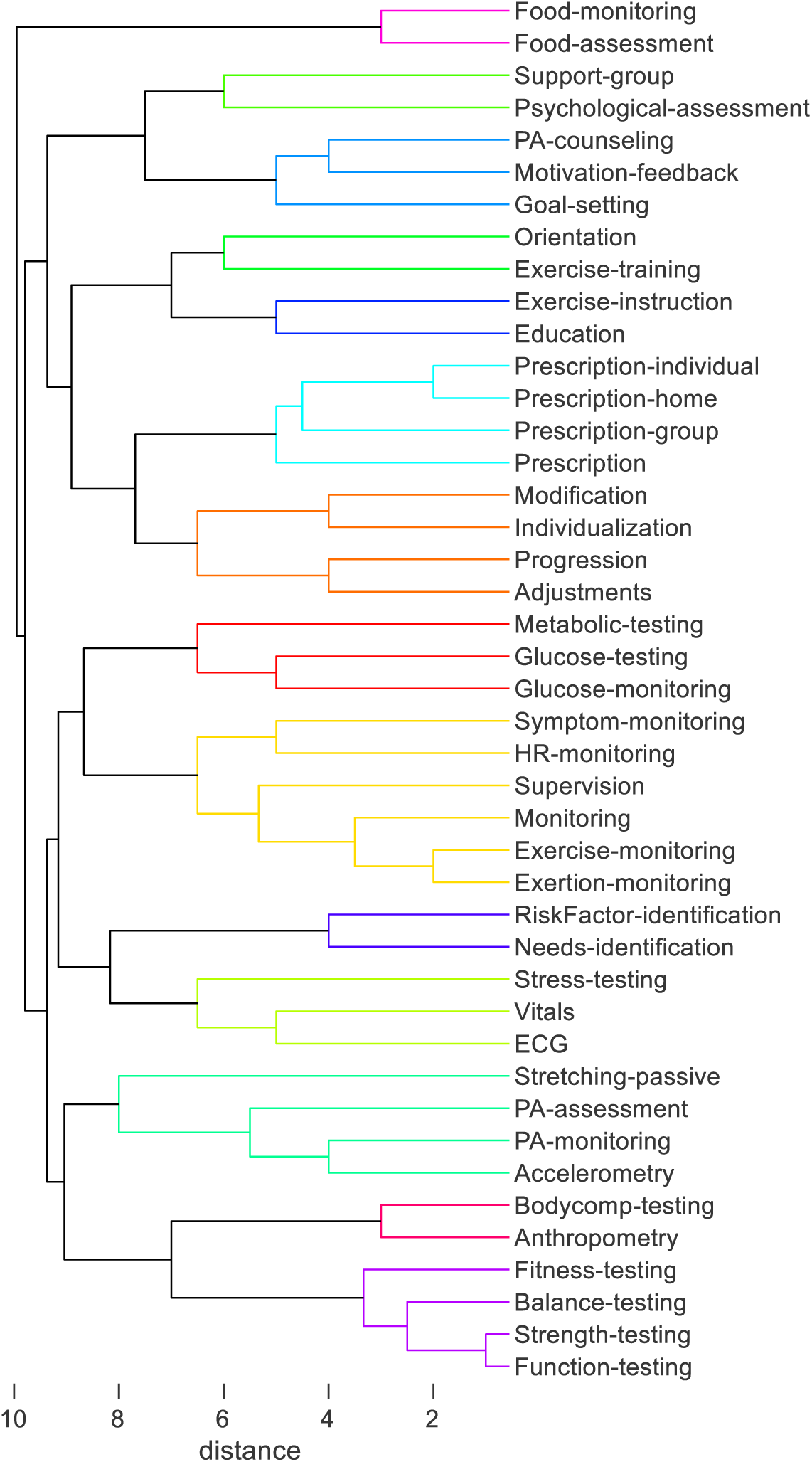
Final hierarchical cluster analysis model approved in the Delphi process

**Supplement 11.**
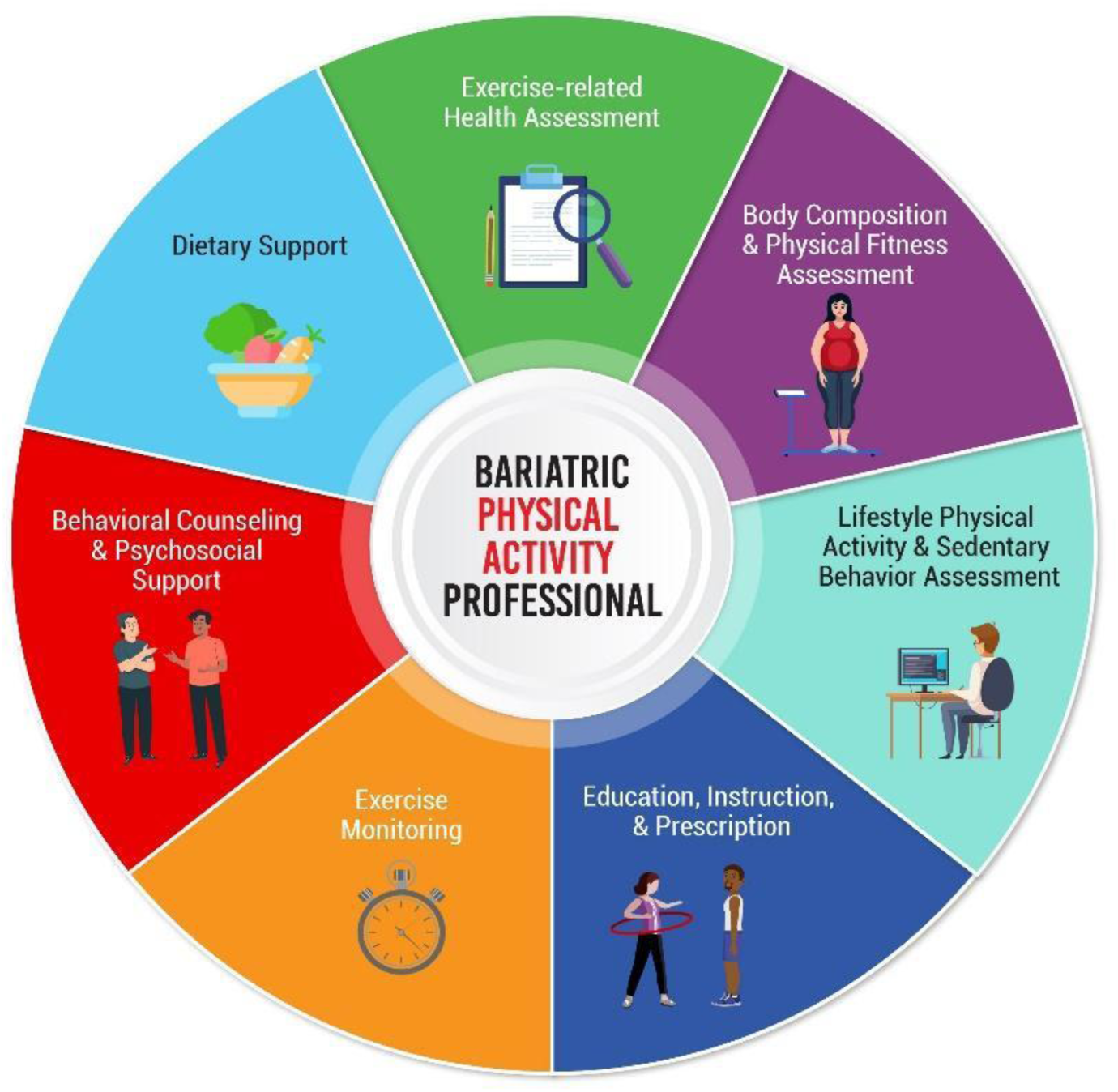
Illustrated final model (Version A) for clinical use. Image by Osama Alowaish, M.S., Teachers College – Columbia University, NY, NY Copyright – The Authors (Stults-Kolehmainen et al., 2023). Use of figure permitted with Authors permission.

## Survey: Issues in Bariatric Exercise

The following statements were rated an average of 9.0, indicating “imperative” was rated by all 10 respondents: 1) “Pre- and post-operative PA/exercise guidelines for metabolic and bariatric (MBS) patients are needed”, 2) “MBS programs need to include PA/exercise as part of multidisciplinary care”. The following questions were rated by USA respondents, also receiving unanimous agreement that they are “imperative”: 1) MBS patients should receive customized and individualized PA/exercise guidance from credentialed exercise professionals with expertise in PA prescription and intervention, 2) Exercise services in MBS settings should be standardized and reimbursed.

## Re-review

The higher order tasks most represented in the literature were “Real-time monitoring, instruction and feedback” at 73%. The task HOT least represented was “stress testing” at 34%. In the review of differences in tasks performed by profession, two higher-order task categories (HOTs) were different. Exercise physiologists were determined to perform both Psychosocial support (HOT 13) and Dietary support (HOT 14) to a greater degree. See Supplement 12.

**Supplement 12a.**
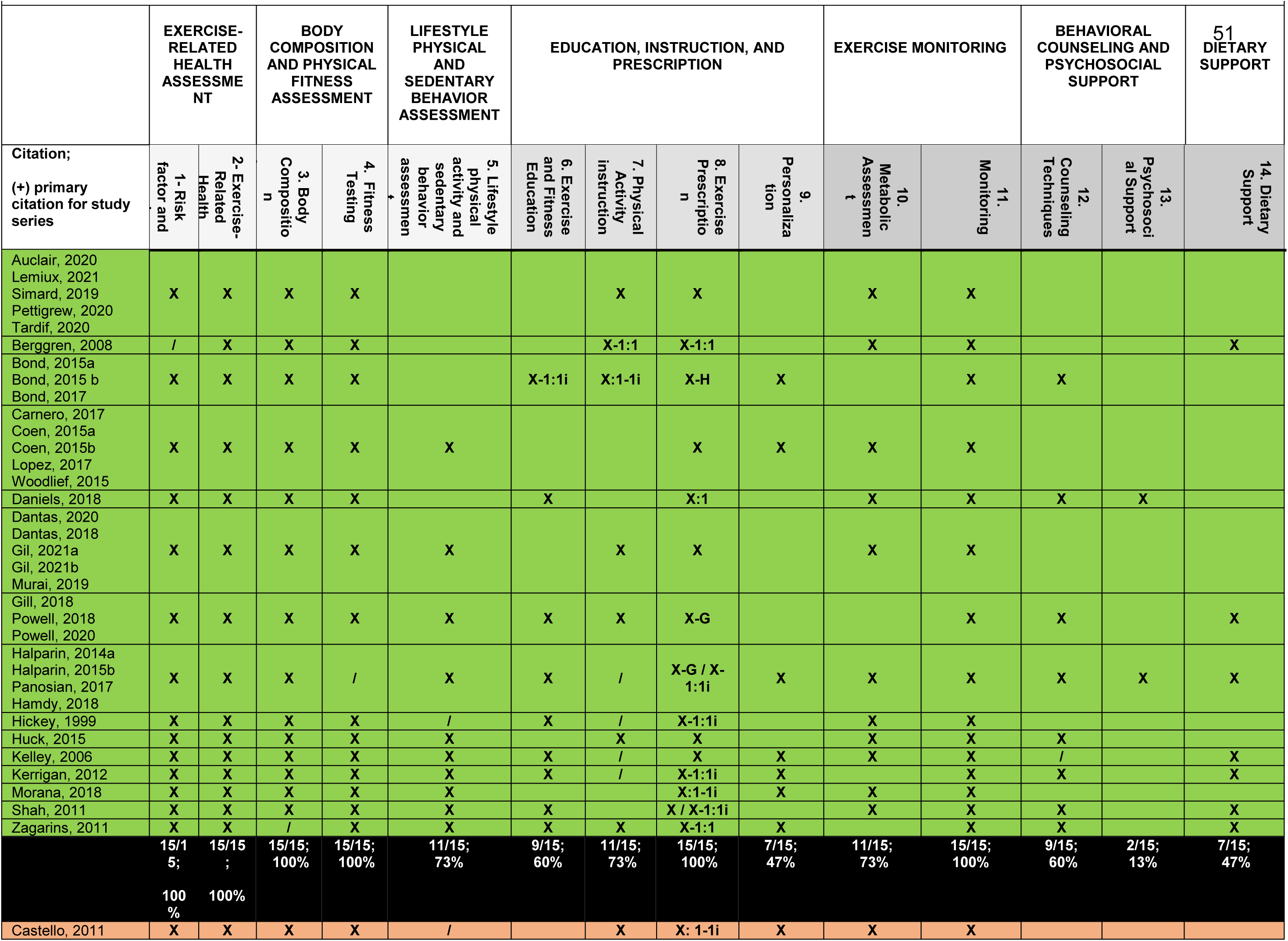
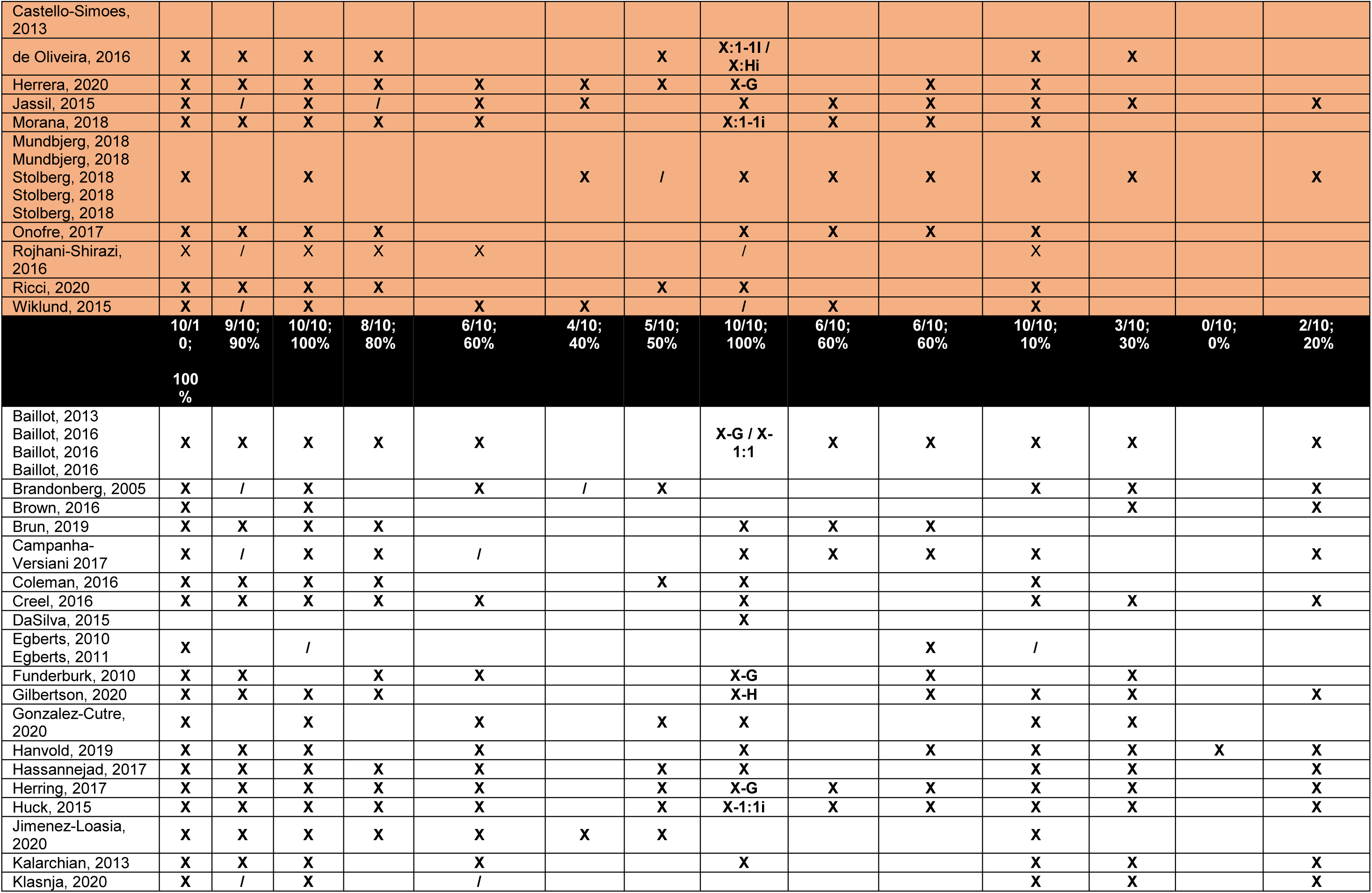
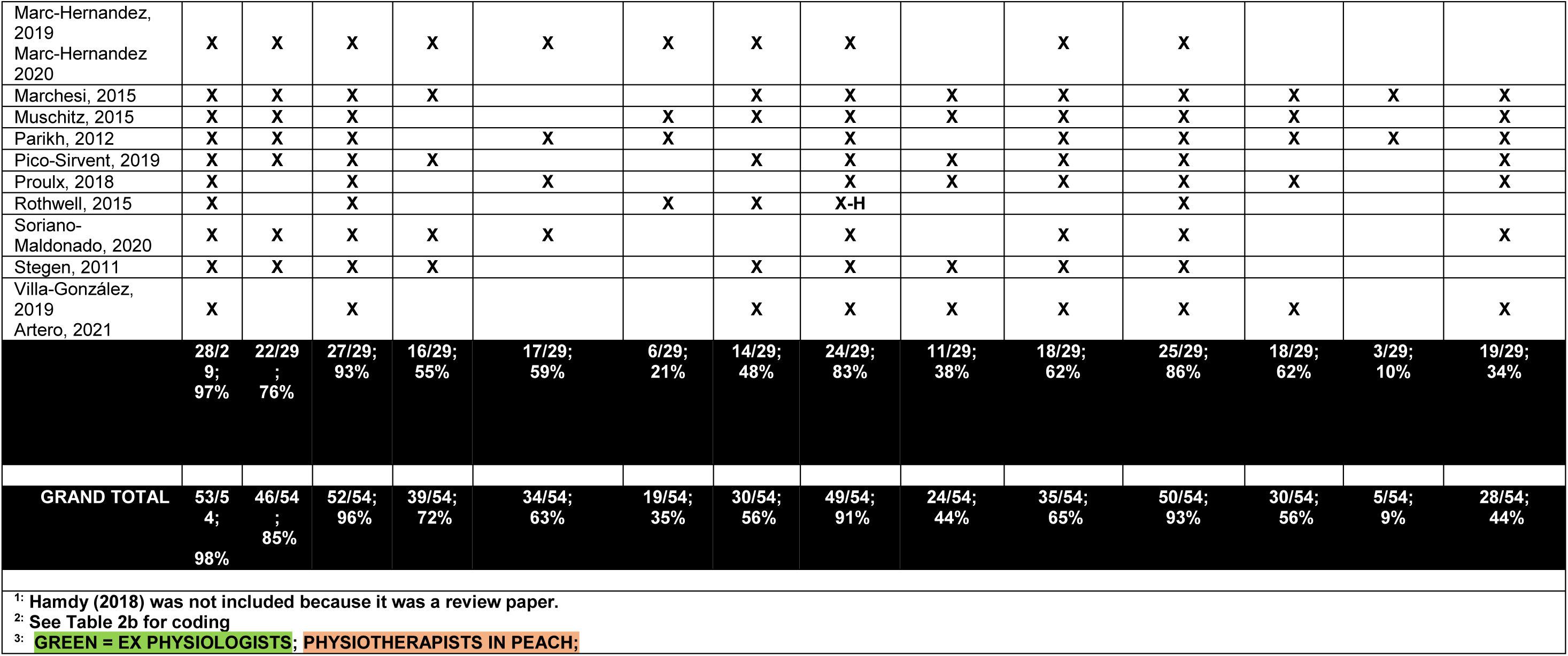
Patient-care-related tasks completed by the exercise professional for each study: Recode and re-review for 14 higher-order themes (HOTs) within 7 super higher-order themes (SHOTs) ^1, 2, 3^.

**Supplement 12b:** Coding for Table 2a.

| **Supplement 12b: Coding for Table 2a** |  |  |  |  |  |
| --- | --- | --- | --- | --- | --- |
|  | X: Explicitly stated<br>/: Implied | Setting | Standardized vs Individualized | Example format | Note |
| X-G | Explicitly stated; | Group setting | Standardized, by definition! | classroom setting; clinic; hospital |  |
| X-1:1 | Explicitly stated; | 1-on-1 training | Standardized | Exam room at clinic, hospital recovery |  |
| X-1:1i | Explicitly stated; | 1-on-1 training | Individualized | Exam room at clinic, hospital recovery |  |
| X-O | Explicitly stated | Online (independent- without the exercise professional present) | Standardized | Video tutorials |  |
| X-Oi | Explicitly stated | Online | Individualized | Interactive/automated digital systems | None of the studies may have this |
| X-H | Explicitly stated | Home (independent- without the exercise professional present) | Standardized | Ex Rx they are given to do at home, same for everyone (similar to InfoSheet, below) |  |
| X-Hi | Explicitly stated | Home (independent- without the exercise professional present) | Individualized | Home Exercise Program (HEP) given at discharge, similar to what physical therapists give |  |
| X-T | Explicitly stated | telephone or video-conferencing | Standardized | Zoom group |  |
| X-Ti | Explicitly stated | telephone or video-conferencing | individualized | Zoom one-on-one |  |
| X-IS | Explicitly stated | Info sheet | Standardized, by definition! | Just informational, not a program, per se, like above. |  |

## DISCUSSION

Increased PA, including exercise, is essential for the long-term success of patients undergoing MBS and helps to maintain health improvements even in the short-to-medium term ^(4–6, 8, 38)^. However, many patients do not become active after surgery. Supports that could offset this inactivity (e.g., PA prescription, counseling, supervised exercise programs) are often not provided as part of clinical care. While there is a clear need for exercise professionals in the multidisciplinary care of patients undergoing MBS, the specifics of their professional background and tasks they perform in a bariatric setting are unclear. To better clarify the roles and rationale for exercise professionals in MBS, we conducted a unique and rigorous multimethod analysis of all exercise programs peri-MBS. We extracted tasks of the exercise professional, submitted these to an expert group of clinicians for sorting into themes, used a sophisticated analysis to create a hierarchical model of these job tasks, and further consulted the expert group for their unique perspectives.

Overall, our study found exercise professionals perform a large set of job tasks related to the clinical care of the patient. Forty-three identified tasks were classified into 14 higher order themes and then seven super-higher order themes. These seven included: (1) Exercise-related health assessment, (2) Body composition and physical fitness assessment, (3) Lifestyle physical activity and sedentary behavior assessment, (4) Education, instruction, and prescription, (5) Exercise monitoring, (6) Behavioral counseling and psychosocial support, and (7) Dietary support. The most common skills reported in the literature were: supervision of exercise, fitness testing, exercise prescription, heart rate monitoring, and physical activity counseling. However, the expert group decided to not rank tasks or their categories by commonality but treat them as relatively equal in importance to avoid overlooking other key functions of the exercise professional. Tasks conducted by exercise professionals in bariatric treatment align closely with similar practice in other clinical settings. For instance, our seven SHOT categories overlapped considerably with core components categories from cardiac rehabilitation (e.g., Psychosocial Management, PA Counseling, Exercise Training Evaluation) ^(39, 40)^ and with performance domains from the American College of Sports Medicine (ACSM) clinical exercise physiology credential (e.g., Exercise Testing, Exercise Prescription, Exercise Training and Leadership, Education and Behavior Change) ^(41)^. The expert group noted that some tasks should not be included, such as post-operative PA clearance (frequently provided by an advanced practice registered nurse [APRN], physician’s assistant or surgeon). The group also indicated it might be worthwhile including other psychosocial support factors as well, including lessening stigmatization, which may contribute to PA/exercise avoidance in bariatric patients ^(42, 43)^. Overall, the results indicate that the exercise professional may have skills and abilities to provide a larger role than that suggested by Mechanick ^(44)^ and others ^(45)^, which appear to limit tasks to “lifestyle medicine evaluation”, physical activity counseling, monitoring adherence to PA guidelines and supervising exercise sessions.

How exercise professionals are classified ostensibly varies by country. Research reviewed in this study, demonstrated the exercise professional was most often an exercise physiologist in the USA, and a physiotherapist/physical therapist outside the USA, the two professions noted by the ASMBS Integrated Health Section ^(46)^. It is possible that physical therapists are frequently utilized due to reimbursement issues, but we were unable to determine this in the current analysis. It is important to note that in Australia, Canada and the United Kingdom, like the USA, exercise physiologists work independent of physical therapists ^(47, 48)^, and in some countries, like Singapore, they may work together. In Brazil, the exercise professional was typically a physical educator. Regardless, our analysis determined that the job tasks exercise professionals conducted in MBS differed on only two dimensions (i.e., Psychosocial support, Dietary support). The expert group did not make any specific recommendations regarding profession, credentialing, or qualifications, but it is clear that credentials of the exercise professional (e.g., exercise physiologist [EP] versus clinical exercise physiologist [CEP]) should match the clinical complexity characteristics of the patients. Moreover, the experts rated as “imperative” that bariatric patients should have customized exercise guidance from credentialed exercise professionals (regardless of whether the professional is an exercise physiologist, physical therapist, or other qualified professional), which and exercise services in the MBS setting should be incorporated into the reimbursable services of care.

The current study has several notable strengths, the first of which is the broad, robust methodological approach used to address the topic, which provided several levels of confirmative evidence for the results. The process was inductive, starting with the literature, and the tasks identified were categorized and refined by an expert group that included a multidisciplinary team of exercise practitioners, scientists, nurses, surgeons, and dietitians from multiple nations. Moreover, the bariatric exercise practitioners involved have several decades of combined experience in this area.

This investigation had some limitations. Studies in the review were predominantly research- and not clinical-focused. As a consequence, most of the patients in these articles were higher functioning – excluding individuals with mobility problems, heart issues, and mental health problems and thus limiting the generalizability of the findings. Thus, one might presume that the literature does not describe a true clinical sample, which may have had implications for the tasks of the exercise professional. To check our model of tasks, a full-scale survey of exercise professionals in MBS across the world should be done as a next step. While highly qualified and experienced, the expert group was a convenience sample, with the implication that there is likely some bias in the analysis. Lastly, a majority of studies provided insufficient reporting of exercise-related tasks. The notable exceptions of this were from a group in Spain ^(49, 50)^ and the USA ^(51)^. However, we aimed to compensate for this by including as many studies as possible on the topic. Furthermore, we contacted numerous authors of the included publications, and their responses are included in these data. Despite these great efforts, it is possible that we missed some important tasks, such as program development or promotion of a physically active environment.

The results of this study have important clinical implications. First, these models will provide a clearer understanding of the role the exercise professional in the MBS setting and the value they bring to a multidisciplinary surgery team ^(46, 52, 53)^. More practically, findings provide practical and useful information that could contribute in four major areas: 1) describing the roles and responsibilities exercise professionals are able to fill, helping create job descriptions, 2) developing CPT/billing codes for reimbursement (aligning with how cardiac rehab was developed), helping to spur other funding for positions, 3) credentialing exercise professionals in bariatric care and development of “core competencies” ^(40)^ and 4) helping to coordinate care from clinical exercise personnel, perhaps in a system of coordinated exercise care. Future needs include PA guidelines for patients undergoing MBS, perhaps similar to those from Exercise and Sports Science Australia ^(3)^ or those recently established for type 2 diabetes. ^(54)^ To this end, it is important to generate more experimental data from well-designed studies on the greater role of PA/exercise in the maintenance of body mass after surgery, as well as other outcomes, and to clarify the ideal type, duration, intensity, progression of such exercise. There also needs to be better reporting of roles and tasks performed by the exercise professional in surgery, as outlined by the CERT guidelines ^(49, 50)^.

Future work should also describe: 1) important knowledge, skills, and competencies (e.g., identifying appropriate exercise protocols based on various health conditions and physical limitations, monitoring blood pressure during exercise and testing, resting and exercise ECG, etc.) ^(55)^, 2) how tasks vary with surgical benchmarks (pre, post and long-term after surgery) and types of programming (e.g., supervised exercise training versus physical activity counseling, 3) scope of practice, and 4) how exercise professionals collaborate as a member of the multidisciplinary team commonly comprised of nurses, dietitians, psychologists, and other providers. For instance, it is likely the case that some tasks described in our model are not solely the responsibility of the exercise professional. More studies are needed to describe the translation and implementation of bariatric and medical weight management PA programs for standardization, reproducibility and to strengthen the case for the role of the exercise professional in MBS and even the greater realm of healthcare.

## CONCLUSION

To our knowledge, this is the first study to systematically investigate the role of the exercise professional in a metabolic and bariatric surgery (MBS) setting. Exercise is regarded as an important facet of multidisciplinary surgical care, but has been poorly implemented, and many programs do not have an exercise professional within their team. It is also important to note that patients want access to exercise professionals before and after their surgery ^(56)^, which, altogether underpins why we completed the first systematic and multimethod investigation of the exercise professional in MBS. We searched for and analyzed every published bariatric exercise/PA program from inception to 2022. Our expert group of bariatric exercise professionals, allied clinicians, and scientists reached a consensus on seven major classifications of patient-centric job tasks for the exercise professional. Tasks varied marginally by the specific field of the exercise professional (e.g., exercise physiologist, physiotherapist/physical therapist, others). Results from our study provide convergent validity with other exercise professional credentialing bodies. We believe that data from this rigorous and unique analysis helps to fill an important gap, enabling continued progress to solidify these positions, while informing new and more precise recommendations on supporting and standardizing comprehensive exercise treatment for obesity. It is also important for governing medical associations across the world to formally recognize experienced exercise professionals as playing pivotal roles in the overall multidisciplinary healthcare for patients undergoing MBS surgery.

## ACKNOWLEDGEMENTS

The authors wish to thank Jessie Moore, R.N., A.P.R.N. (Yale New Haven Hospital), Jennifer Guan, D.P.T., Edd Taylor, Ph.D. (University of Colorado) and Amanda Divin, Ph.D. (Texas A&M University - Commerce) for thoughtful critiques and technical assistance. Rebecca Lamberti, R.N., B.S.N., M.S.N., D.N.P. (Yale New Haven Hospital) and Diana Rivera, R.N., B.S.N., C.N.M.L. (Yale New Haven Hospital) provided strategic administrative and logistical support. Raw data is available upon reasonable request to the corresponding author.

Laura Richardson is past president of the Clinical Exercise Physiology Association (CEPA) and is credentialed as a registered clinical exercise physiologist (RCEP) and ACSM-CEP. Carol Garber Ewing is a past president and current Treasurer of the ACSM and is credentialed as a RCEP and ACSM-CEP. John Morton is the past president of the American Society of Metabolic and Bariatric Surgery (ASMBS) and is the present chair of the American College of Surgeons, Metabolic and Bariatric Surgery Accreditation and Quality Improvement Program. Saber Ghiassi is the current president of the Connecticut Chapter of ASMBS. Janet Huehls is credentialed as RCEP and ACSM-CEP. Matthew Stults-Kolehmainen, Susannah Williamson, and Nina Brojan Heyman are credentialed by the ACSM as certified exercise physiologists (ACSM-EP). Dale Bond is associate editor for the Integrative Health section of SOARD.

## COMPETING INTERESTS

None

## AUTHOR CONTRIBUTIONS

The concept was developed by MSK, DB, SG, and BG. MSK, DB, LR and LH formed the Core Group. The Expert Group and advisory panel consisted of MSK, LR, JH, CEG, MD, EB, BM, AD, DB, LH, BG, ERM, GA, JM, OA, SW and SG. The literature review was conducted by MSK. Data extraction was conducted by MSK, MD, OA, and SW. CA conducted the statistical modelling. Statistical consulting was provided by MH. MSK, DB, BG, LR, and LH wrote the manuscript. SB created the artwork. OA provided additional illustration support. NBH was the study auditor. All authors read and critiqued the paper and provided final approval of the manuscript.

## FUNDING

Bruno Gualano is supported by FAPESP, CNPq and CAPES. LH is supported by the National Institute for Health and Care Research (NIHR) - Leicester Biomedical Research Centre (BRC). [The views expressed are those of the author(s) and not necessarily those of the NIHR or the Department of Health and Social Care.] GA was supported by American Heart Association Grant #852679 (GA, 2021–2024), and the National Institute of Diabetes, Digestive, and Kidney Diseases of the National Institutes of Health under a mentored research scientist development award (K01DK129441).

## DATA AVAILABILITY STATEMENT

All data produced in the present study are available upon reasonable request to the authors.

## Notes

### Competing Interest Statement

The authors have declared no competing interest.

### Summary of Updates

There are minor, but substantive, changes in this revision. Supplements have been reordered. One supplement was changed to a table. The discussion was expanded.

